# Integrated Analyses of Growth Differentiation Factor-15 Concentration and Cardiometabolic Diseases in Humans

**DOI:** 10.1101/2021.12.15.21267719

**Authors:** Susanna Lemmelä, Eleanor M. Wigmore, Christian Benner, Aki Havulinna, Rachel M. Y. Ong, Tibor Kempf, Kai C. Wollert, Stefan Blankenberg, Tanja Zeller, James E. Peters, Veikko Salomaa, Maria Fritsch, Ruth March, Aarno Palotie, Mark Daly, Adam S. Butterworth, Mervi Kinnunen, Dirk S. Paul, Athena Matakidou

## Abstract

Growth differentiation factor 15 (GDF15) is a stress response cytokine that is elevated in several cardiometabolic diseases and has attracted interest as a potential therapeutic target. To further explore the association of GDF15 with human disease, we conducted a broad study into the phenotypic and genetic correlates of GDF15 concentration in up to 14,099 individuals. Assessment of 772 traits across 6,610 participants in FINRISK identified associations of GDF15 concentration with a range of phenotypes including all-cause mortality, cardiometabolic disease, respiratory diseases and psychiatric disorders as well as inflammatory markers. A meta-analysis of genome-wide association studies (GWAS) of GDF15 concentration across 3 different assay platforms (n=14,099) confirmed significant heterogeneity due to a common missense variant rs1058587 in *GDF15*, potentially due to epitope-binding artefacts. After conditioning on rs1058587, statistical fine-mapping identified 4 independent putative causal signals at the locus. Mendelian randomisation (MR) analysis did not find evidence of a causal relationship between GDF15 concentration and cardiometabolic traits. Using reverse MR, we identified a potential causal association of body mass index on GDF15 (IVW *p*_FDR_=0.0072). Taken together, our data do not support a role for elevated GDF15 concentrations as a causal factor in human cardiometabolic disease but support its role as a biomarker of metabolic stress.

## Introduction

Obesity accounts for an estimated 2.8 million deaths worldwide and 2.3% of global disability-adjusted life years (Organisaton.). Obesity has been causally linked to a variety of cardiometabolic risk factors and diseases, including fasting insulin, systolic blood pressure and type 2 diabetes (Holmes et al., 2014). Therefore, interventions to reduce obesity are recommended in the management of these diseases (Yumuk et al., 2016). Currently, the most effective obesity intervention is bariatric surgery (Jammah, 2015); however, its clinical utility is limited as it is highly invasive and accompanied by a high risk of long-term complications such as gastroesophageal reflux disease and nutritional deficiencies (Mesureur & Arvanitakis, 2017). New therapies such as GLP-1 receptor agonists aim to target obesity via appetite suppression but are associated with common side effects including nausea and diarrhoea (Wilding et al., 2021). The development of additional therapeutic strategies targeting obesity as an underlying cause and pathology of cardiometabolic disease represents an area of unmet clinical need.

GDF15, a distant member of the transforming growth factor-β family (Bootcov et al., 1997; Emmerson, Duffin, Chintharlapalli, & Wu, 2018), is upregulated in response to cellular stress and several diseases, including cancer. It has been suggested that GDF15 functions as a stress response agent implicated in organ injury (Kempf et al., 2006; V. W. W. Tsai, Husaini, Sainsbury, Brown, & Breit, 2018; Zimmers et al., 2005). GDF15 was originally identified to play a role in tumour-induced anorexia/cachexia in mice through appetite regulation (Johnen et al., 2007). The body weight reduction is considered to be mainly driven by food intake inhibition mediated by its receptor GDNF-family receptor alpha like (GFRAL) in distinct areas of the brainstem (Emmerson et al., 2017; Hsu et al., 2017; Mullican et al., 2017; Yang et al., 2017). GDF15 plasma levels are elevated in animal models of obesity (Lockhart, Saudek, & O’Rahilly, 2020; Xiong et al., 2017), and administration of recombinant protein robustly lowers body weight in obese and diabetic animals, including non-human primates (Mullican et al., 2017; Xiong et al., 2017). The beneficial effects on glycemic control is considered to be mediated mainly by the body weight loss. Genetic overexpression of Gdf15 results in decreased body weight and increased resistance to obesity associated with high-fat diets, whereas Gdf15 and Gfral deficient mice are more susceptible to diet-induced obesity (Mullican et al., 2017; O’Rahilly, 2017; Tran, Yang, Gardner, & Xiong, 2018). Recent preclinical work has identified GDF15’s role as a sentinel protein, upregulated in response to various ingested toxins triggering nausea-related behaviour in rodents assessed by pica and CTA (Conditioned Taste Aversion) (Borner, Wald, et al., 2020; Patel et al., 2019).

There is a lack of data supporting the treatment effect of GDF15 in humans. However, observational studies have revealed strong positive correlations of GDF15 plasma levels with body mass index (BMI), insulin resistance, age and mean arterial blood pressure (in obese individuals) (V. W. Tsai et al., 2015; Vila et al., 2011). Higher GDF15 levels have also been associated with all-cause mortality as well as mortality associated with heart failure and acute myocardial infarction, cancer, advanced heart failure and end-stage chronic kidney disease (Adela & Banerjee, 2015; Khan et al., 2009; Nair et al., 2017; Wiklund et al., 2010). Metformin therapy, a type 2 diabetes treatment, has been shown to depend on GDF15 to lower body weight in mice and plasma GDF15 levels increased up to 40% upon metformin therapy in humans (Coll et al., 2020; Day et al., 2019). Recent data suggest that GDF15 is involved in hyperemesis gravidarum (Fejzo, Arzy, Tian, MacGibbon, & Mullin, 2018), supporting the hypothesis that GDF15 triggers anorexia and subsequent weight loss, at least partly, through the induction of malaise (Borner, Shaulson, et al., 2020). Mendelian randomisation (MR) has previously been applied to explore the causal relationship of GDF15 levels and cardiometabolic diseases, with causal associations reported for high-density lipoprotein (HDL) cholesterol and bone mineral density (BMD) (Cheung, Tan, Au, Li, & Cheung, 2019; Folkersen et al., 2020). However, these analyses were based on small genetic studies for GDF15 and have not been replicated.

In this study, we used data from several large biobanks to conduct a systematic and extensive phenotypic and genotypic analysis of GDF15 with cardiometabolic traits and diseases, and to ascertain the causal relationship between GDF15 levels and cardiometabolic traits using MR and protein-truncating variant (PTV) analysis.

## Results

### Association of GDF15 plasma levels with 676 disease outcomes

To systematically assess the relationship between GDF15 plasma levels and clinical phenotypes, we utilised a large Finnish biobank, FINRISK. The FINRISK cohort comprises a cross-sectional population survey carried out over a 40-year period in Finland. GDF15 plasma levels were available for 6,610 participants from the 1997 recruitment cohort linked to 676 disease outcomes and were included in this analysis.

The median GDF15 concentration (measured using an immunoluminometric assay) was 796ng/L (interquartile range, IQR= 664-986) (Supplementary Figure S1). First, we examined the association of baseline characteristics with GDF15 plasma levels (Supplementary Table S1). Age explained 28% of the observed GDF15 variance, with current smoking and BMI accounting for 1.9% and 0.08% of the variance, respectively (Supplementary Table S2). Gender did not show a significant association. All subsequent analyses have been corrected for these covariates (age, gender, smoking and BMI). Analyses corrected for age and gender alone are also presented for comparison (Supplementary Table S3 and S4), although results were similar.

We then investigated potential associations of GDF15 plasma levels with a range of disease phenotypes (both prevalent and incident), focusing on the phenotypes defined by the FinnGen consortium (T, Lahtela E, Havulinna AS, & Consortium., 2020), as these endpoints have undergone additional clinical validation (see Methods). A total of 676 disease endpoints were examined as dependent variables in association analyses with GDF15 plasma levels (the independent variable). After multiple-testing correction using false discovery rate (FDR, P_FDR_ < 0.05), GDF15 was significantly associated with 80 disease endpoints (Table 1 and Supplementary Table S5).

**Table 1.** Disease endpoints associated (*p*_FDR_<1×10^−5^) with GDF15 plasma levels in FINRISK.

| **Disease Endpoint** | **Cases / Controls** | **OR (95% CI)** | **$p_{\text{FDR}}$** |
| --- | --- | --- | --- |
| All-cause mortality | 1057/5481 | 1.79 (1.68-1.90) | $7.5 \times 10^{-24}$ |
| Death due to cardiac causes | 471/6067 | 1.76 (1.61-1.90) | $3.0 \times 10^{-11}$ |
| Atherosclerosis, excluding cerebral, coronary and PAD | 379/6159 | 1.67 (1.51-1.82) | $3.2 \times 10^{-8}$ |
| Diabetes mellitus type 2 | 567/5971 | 1.48 (1.36-1.60) | $3.2 \times 10^{-8}$ |
| Diabetes mellitus | 592/5946 | 1.44 (1.33-1.56) | $9.8 \times 10^{-8}$ |
| Diseases of arteries, arterioles and capillaries | 505/6033 | 1.48 (1.35-1.61) | $3.6 \times 10^{-7}$ |
| Other chronic obstructive pulmonary disease | 221/6317 | 1.84 (1.63-2.04) | $5.7 \times 10^{-7}$ |
| Chronic obstructive pulmonary disease | 235/6303 | 1.77 (1.57-1.97) | $9.6 \times 10^{-7}$ |
| Pneumonia (excl. viral and due to other infectious organisms not elsewhere classified) | 779/5759 | 1.34 (1.24-1.44) | $9.6 \times 10^{-7}$ |
| Type 2 diabetes without complications | 485/6053 | 1.44 (1.31-1.57) | $1.2 \times 10^{-6}$ |
| All pneumonia | 789/5749 | 1.33 (1.23-1.43) | $1.9 \times 10^{-6}$ |
| Chronic kidney disease | 66/6472 | 2.46 (2.14-2.79) | $4.4 \times 10^{-6}$ |
| Influenza and pneumonia | 833/5795 | 1.30 (1.21-1.40) | $4.4 \times 10^{-6}$ |
| Type 2 diabetes with renal complications | 35/6503 | 2.97 (2.57-3.38) | $8.1 \times 10^{-6}$ |
| Sequelae of cerebrovascular disease | 209/6329 | 1.67 (1.47-1.86) | $9.2 \times 10^{-6}$ |
| Alcoholic liver disease | 56/6482 | 2.26 (1.95-2.57) | $9.2 \times 10^{-6}$ |

Results are adjusted for age, gender, smoking and BMI. Abbreviations: OR, odds ratio; CI, confidence interval; COPD, chronic obstructive pulmonary disease; PAD, peripheral artery disease; SAH, subarachnoid haemorrhage. ICD codes for these disease endpoints have been published (T et al., 2020).

The most significant clinical associations observed were with all-cause mortality (logistic regression odds ratio, OR=1.8, CI=1.7 – 1.9, *p-*value=9.8×10^−27^) and mortality due to cardiac diseases (OR=1.8, CI=1.6 – 1.9, *p-*value=7.7×10^−14^). We also observed significant associations between GDF15 levels and type 2 diabetes (OR=1.5, CI=1.3 – 1.6, *p-*value=2.2×10^−9^). Further, GDF15 plasma levels were significantly associated with cardiovascular diseases (OR=1.2, CI=1.1–1.2, *p-*value=5.6×10^−6^), as well as subtype endpoints such as atherosclerosis, hypertension, peripheral artery disease and stroke, and chronic kidney disease (OR=2.46, CI=2.1–2.8, *p*-value=4.4×10^−6^) (Table 1). Additional positive associations were observed with respiratory diseases, such as chronic obstructive pulmonary disease (COPD) and pneumonia, and psychiatric disorders, including schizophrenia and mental and behavioural disorders due to psychoactive substance use. Cancer phenotypes were also associated with GDF15 levels, including malignant neoplasm of the respiratory system and intrathoracic organs (OR=1.6, CI=1.3–1.8, *p-*value=0.0015) and malignant neoplasm of the bronchus and lung (OR=1.6, CI=1.3–1.9, *p*-value=0.0029).

### GDF15 plasma concentration associates with prevalent and incident type 2 diabetes and independently predicts all-cause mortality and cardiometabolic diseases

To determine the prognostic potential of GDF15 in predicting incident diabetes, we investigated its plasma levels in relation to prevalent and incident type 2 diabetes in the FINRISK cohort. Type 2 diabetics (prevalent cases) had a 1.3-fold increase in median GDF15 levels compared to non-diabetic controls (median difference=222ng/L). Furthermore, GDF15 levels were significantly higher in prevalent diabetics (diagnosed at plasma sampling, n=37; median=1,204ng/L) compared to both incident cases (diagnosed after plasma sampling, n=500; median= 992ng/L; Wilcoxon *p-*value=0.008) and non-diabetic controls (n=6003; 784ng/L; Wilcoxon *p-*value=5.2×10^−11^) (Supplementary Figure S2). GDF15 plasma levels were also significantly higher in incident cases than in non-diabetic controls (Wilcoxon *p-*value=6.2×10^−51^). Together, these data indicate that increased GDF15 plasma levels could represent an early biomarker in pre-diabetic individuals.

Next, we carried out a Cox regression survival analysis on all-cause mortality, type 2 diabetes and cardiovascular disease. During a 10-year follow-up period, 393 (6%) study subjects had died, 97 (2%) developed diabetes and 438 (7%) developed cardiovascular disease. Models accounted for blood pressure medication, smoking, total cholesterol, HDL cholesterol, BMI, prevalent cardiovascular disease and mean systolic blood pressure. Our results revealed that GDF15 plasma concentration is an independent predictor of all-cause mortality (hazard ratio, HR=1.7, *p-*value=9.2×10^−12^), type 2 diabetes (HR=1.40, *p-*value=0.02) and cardiovascular disease (HR=1.4, *p-*value=1.5×10^−6^, Figure 1, Supplementary Table S6 and Figure S3). Individuals with GDF15 plasma levels above 967ng/L (the upper quartile) were more than twice as likely to develop type 2 diabetes compared to individuals with lower levels (HR=2.2, CI=1.4–3.5, *p-*value=7.1×10^−4^).

**Figure 1.**
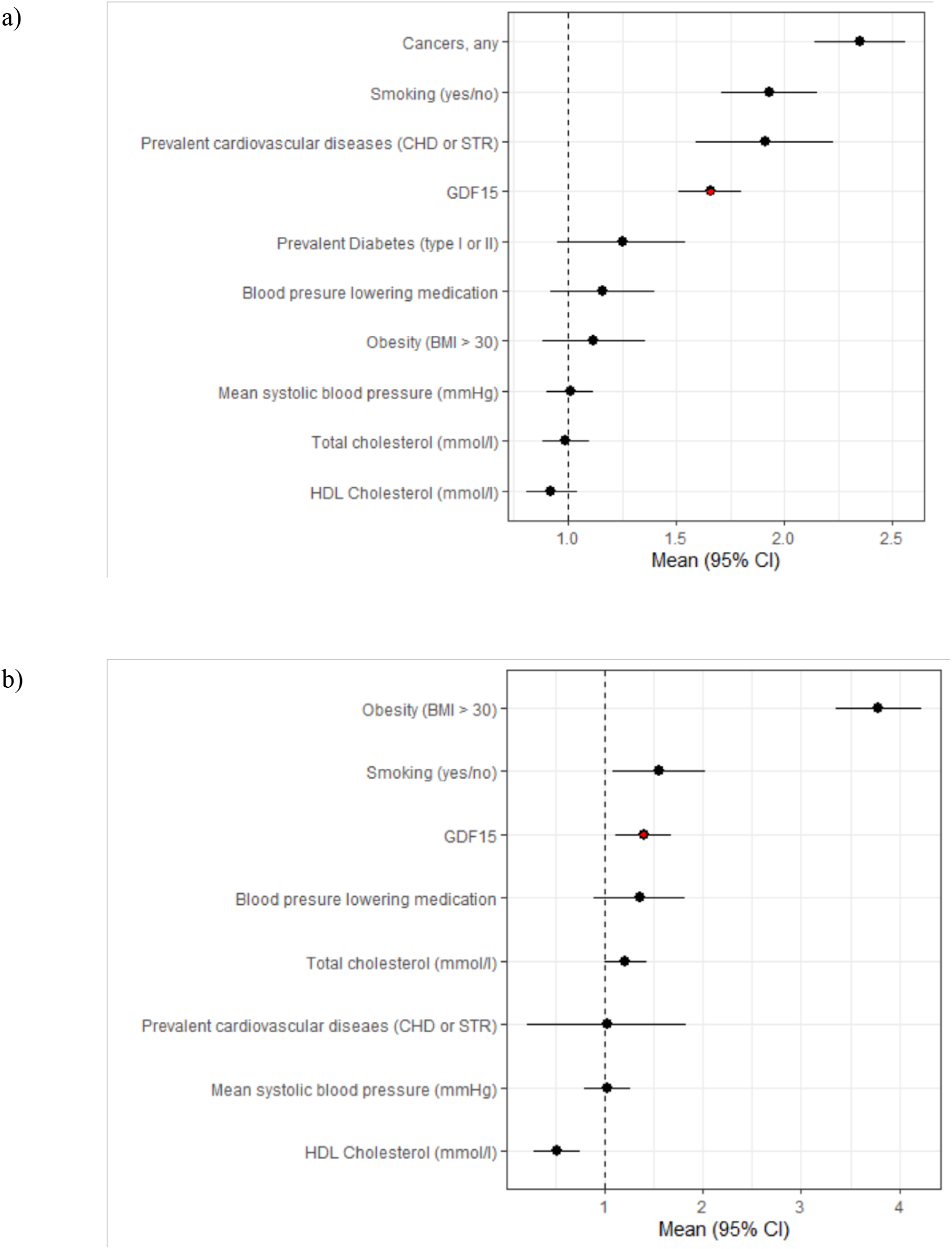
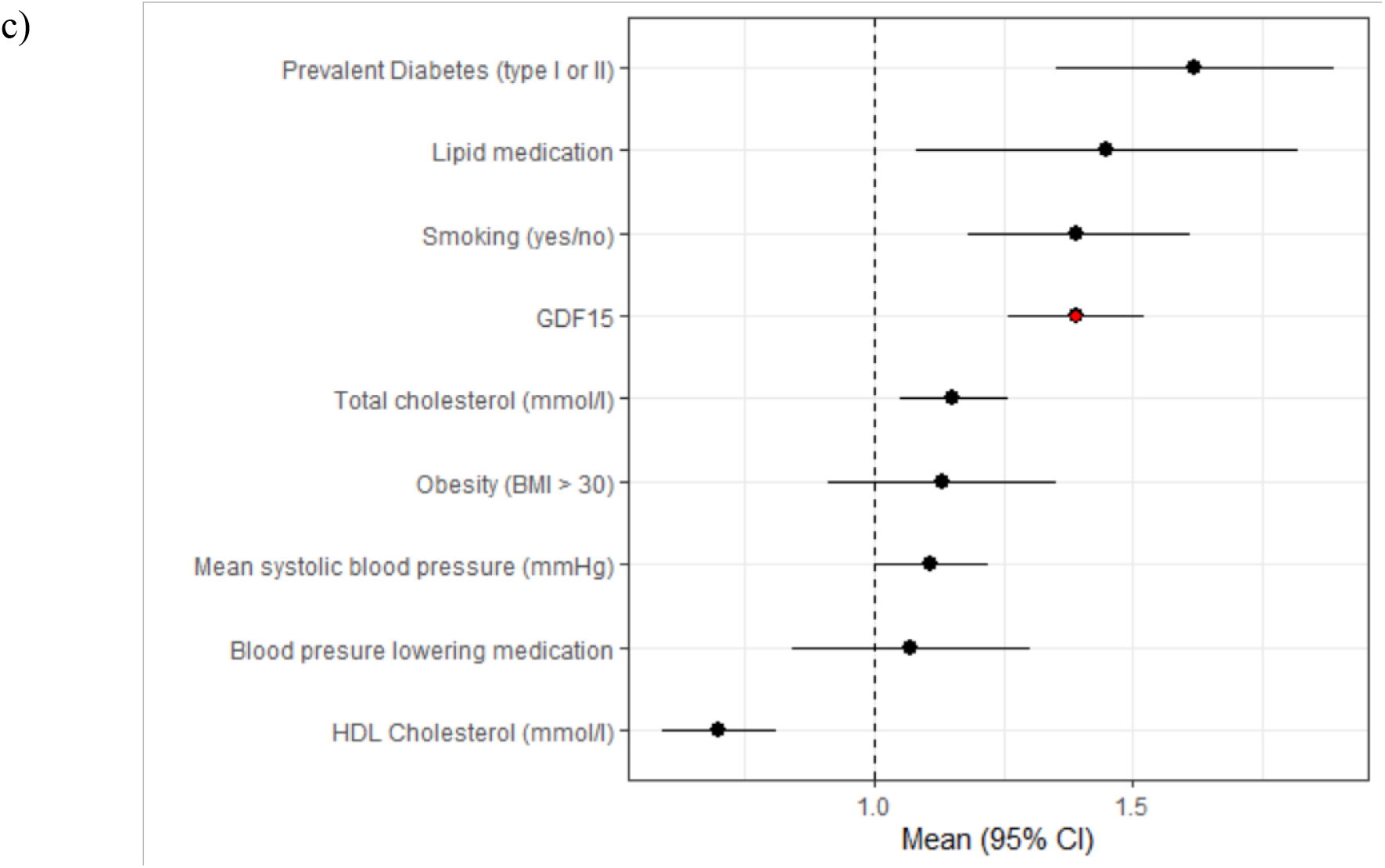
Forest plots of Cox proportional hazard for independent predictors of (a) all-cause mortality, (b) cardiovascular disease and (c) diabetes. GDF15 is highlighted in red and variables are ordered by highest hazards ratio. Abbreviations: BMI, body mass index; GDF15, growth differentiation factor-15; CHD, coronary heart disease; STR, stroke; HDL, high density lipoprotein.

### Association of GDF15 plasma levels with 96 quantitative biomarkers

We then performed an association analysis of GDF15 plasma levels with 96 quantitative biomarkers in FINRISK. After multiple-testing correction (P_FDR_<0.05), significant associations were observed with 45 biomarkers, most notably, with blood markers known to be associated with inflammation (Supplementary Table S7).

The most significantly associated biomarkers were mid-regional pro-adrenomedullin (MR-proADM, beta=0.24, *p-*value=1.2×10^−91^), C-reactive protein (CRP, beta=0.22, *p-*value=4.6×10^−64^) and hepatocyte growth factor (HGF, beta=0.17, *p-*value=4.2×10^−43^). All three markers have been previously reported to be non-specifically elevated in a number of human diseases and have known prognostic value (Jackson et al., 2016; Madonna, Cevik, Nasser, & De Caterina, 2012; Matsumoto, Umitsu, De Silva, Roy, & Bottaro, 2017; Miller, Redfield, & McConnell, 2007; Peacock, 2014). Significant associations were also observed between GDF15 and components of metabolic syndrome, specifically, levels of triglycerides (beta=0.13, *p-*value=2.4×10^−22^), fasting insulin (beta=0.10, *p-*value=1.2×10^−13^) and waist-to-hip ratio (beta= 0.049, *p-*value=7.0×10^−10^).

### Genome-wide association studies of plasma levels of GDF15 quantified using 3 different assays in 2 independent cohorts

We performed genome-wide association studies (GWAS) to identify the genetic determinants of GDF15 plasma levels using two independent cohorts (FINRISK and INTERVAL) and three GDF15 quantification methodologies (immunoluminometric assay in FINRISK and Olink and SomaScan proteomic assays in INTERVAL; see Supplementary Materials). Results from individual GWAS analyses are reported in Supplementary Tables S8-10. We then performed a meta-analysis of the genome-wide significant variants identified in the GWAS performed in the FINRISK, INTERVAL-SomaScan and INTERVAL-Olink cohorts to generate a combined statistic and heterogeneity value. The analysis identified four genetic variants (rs1059369, rs1054221, rs1227734, rs189593084) at the *GDF15* locus to be associated with GDF15 plasma levels (*p*-value<5×10^−8^) (Supplementary Table S11).

Two variants had a strengthened signal in the combined meta-analysis, rs1054221 and rs1227734 (Supplementary Table S11), However, five genetic associations, which were genome-wide significant for two of the three GWAS, displayed substantial heterogeneity, e.g. rs16982345 (heterogeneity I^2^=99.8%, heterogeneity *p*-value=7.6×10^−180^; Supplementary Table S11). By exploring the LD structure between the heterogeneous variants (Supplementary Table S12), we found that these variants were all located within the same LD block that included the missense variant rs1058587 (p.H202D). Heterogeneity in GWAS results of GDF15 levels has been observed in other studies (Jiang et al., 2018; Pietzner et al., 2020) and significant variations in GDF15 antibody binding affinity dependent on rs1058587 have been previously reported (Fairlie et al., 2001).

### Mitigation of epitope binding effects due to the rs1058587 missense variant

To remove any potential epitope effects, we applied conditional analysis on rs1058587 when linking plasma GDF15 concentrations with disease phenotypes and quantitative traits. A total of 59 significant disease associations and 43 biomarker associations were identified. These associations largely did not differ from the unconditioned results, although 21 fewer disease associations reached significance (Supplementary Table S13-14).

To minimize confounding of the GWAS results from rs1058587 epitope effects, we conditioned the association signal on chromosome 19 on rs1058587 in both FINRISK (Supplementary Table S15) and INTERVAL-SomaScan (Supplementary Table S16). INTERVAL-Olink did not have a significant association at this LD block. We then meta-analysed these conditioned GWAS. Manhattan and Quantile-Quantile (QQ) plots of this GWAS meta-analysis are shown in Table 2, Supplementary Figure S4 and Supplementary Table S15. We found 146 significant associations in this region, with rs1227734 identified as the strongest association (beta=0.50, *p*-value=2.5×10^−187^).

**Table 2.**
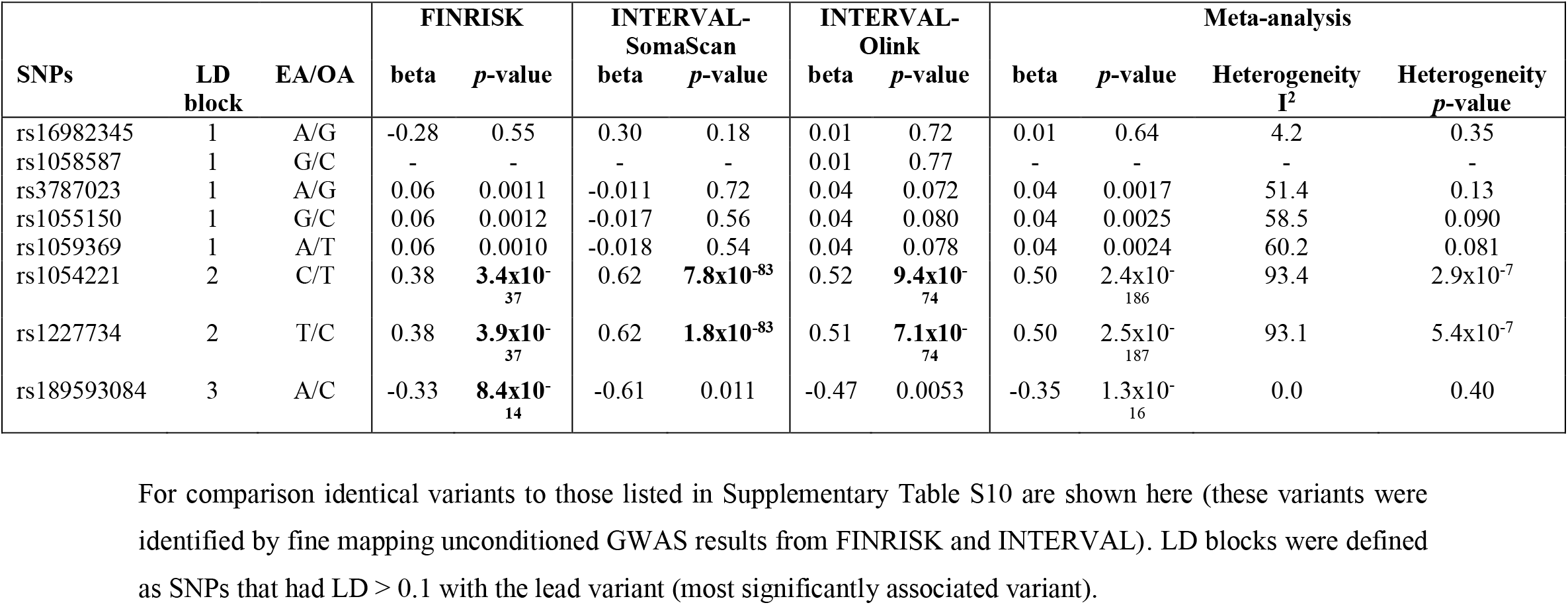
Meta-analysis of GDF15 GWAS conditioned on rs1058587 in FINRISK and INTERVAL.

| SNPs | LD block | EA/OA | FINRISK |  | INTERVAL-SomaScan |  | INTERVAL-Olink |  | Meta-analysis |  |  |  |
| --- | --- | --- | --- | --- | --- | --- | --- | --- | --- | --- | --- | --- |
|  |  |  | beta | p-value | beta | p-value | beta | p-value | beta | p-value | Heterogeneity I<sup>2</sup> | Heterogeneity p-value |
| rs16982345 | 1 | A/G | -0.28 | 0.55 | 0.30 | 0.18 | 0.01 | 0.72 | 0.01 | 0.64 | 4.2 | 0.35 |
| rs1058587 | 1 | G/C | - | - | - | - | 0.01 | 0.77 | - | - | - | - |
| rs3787023 | 1 | A/G | 0.06 | 0.0011 | -0.011 | 0.72 | 0.04 | 0.072 | 0.04 | 0.0017 | 51.4 | 0.13 |
| rs1055150 | 1 | G/C | 0.06 | 0.0012 | -0.017 | 0.56 | 0.04 | 0.080 | 0.04 | 0.0025 | 58.5 | 0.090 |
| rs1059369 | 1 | A/T | 0.06 | 0.0010 | -0.018 | 0.54 | 0.04 | 0.078 | 0.04 | 0.0024 | 60.2 | 0.081 |
| rs1054221 | 2 | C/T | 0.38 | **3.4x10<sup>-37</sup>** | 0.62 | **7.8x10<sup>-83</sup>** | 0.52 | **9.4x10<sup>-74</sup>** | 0.50 | 2.4x10<sup>-186</sup> | 93.4 | 2.9x10<sup>-7</sup> |
| rs1227734 | 2 | T/C | 0.38 | **3.9x10<sup>-37</sup>** | 0.62 | **1.8x10<sup>-83</sup>** | 0.51 | **7.1x10<sup>-74</sup>** | 0.50 | 2.5x10<sup>-187</sup> | 93.1 | 5.4x10<sup>-7</sup> |
| rs189593084 | 3 | A/C | -0.33 | **8.4x10<sup>-14</sup>** | -0.61 | 0.011 | -0.47 | 0.0053 | -0.35 | 1.3x10<sup>-16</sup> | 0.0 | 0.40 |

Fine-mapping this conditioned meta-analysis revealed four association signals with a causality probability of 0.86. The top configuration consisted of variants rs1054221 (beta=-0.50, *p*-value=2.4×10^−184^), rs3787023 (beta=0.04, *p*-value=0.0017), rs138515339 (beta=-0.27, *p*-value=3.2×10^−11^) and rs141542836 (beta=0.61, *p*-value=1.9×10^−40^) and had a regional heritability of 7%. Functional annotation and eQTL information can be found in Supplementary Table S18 and the Supplementary materials, respectively.

### Mendelian randomisation analysis reveals no causal relationship between GDF15 plasma levels and cardiometabolic traits

Observed associations of GDF15 plasma levels with obesity-related diseases have been presented here and found in previous reports (V. W. Tsai et al., 2015; Vila et al., 2011) and variants in the *GDF15* gene region have been associated with cardiovascular traits, cholesterol, waist-hip-ratio (WHR) and BMI (Wang et al., 2021). We therefore applied MR to assess the relationship between genetically-determined GDF15 plasma levels and BMI, WHR, glucose and type 2 diabetes. We also included HDL cholesterol and BMD as additional outcomes in our MR analysis due to the positive MR results in previous studies (Cheung et al., 2019; Folkersen et al., 2020).

We applied two-sample MR (Bowden, Davey Smith, & Burgess, 2015) using LD clumping (R^2^ < 0.1, within 1.5Mb of either side of the lead SNP) and identified eight independent genetic instruments from the conditioned meta-analysis on chromosome 19: rs1227734, rs62122429, rs1059022, rs181004295, rs75119307, rs11673678, rs138185133 and rs73923175 (regional heritability=0.071). Association statistics between the conditioned instrumental variables (IVs) and exposure (GDF15) were taken from the above meta-analysis, and the association statistics between the IVs and outcome were extracted from publicly available GWAS summary statistics (see Methods). These values were applied to a fixed effect inverse variance weighted (IVW) MR, as well as MR-Egger and MR-PRESSO methods (Verbanck, Chen, Neale, & Do, 2018), in order to detect horizontal pleiotropy. MR using the conditioned variants did not find significant associations of genetically-determined GDF15 plasma levels with any of the examined traits. A nominal association was found with WHR but this did not pass multiple-testing correction (Table 3). Horizontal pleiotropy was detected in the association of genetically-determined GDF15 plasma levels with BMI (Global pleiotropy test *p*-value=0.012), HDL cholesterol (Global pleiotropy test *p*-value=0.021) and BMD (Global pleiotropy test *p*-value=0.010) using MR-PRESSO but not MR-Egger. We did not replicate findings of causality of genetically-determined GDF15 plasma levels with BMD and HDL cholesterol, despite two independent studies suggesting otherwise (Cheung et al., 2019; Folkersen et al., 2020).

**Table 3.** Mendelian randomisation results for exposure genetically-determined GDF15 plasma levels with cardiometabolic outcomes.

| Methods | IVW (fixed) |  | MR-Egger |  |  | MR-PRESSO |  |  |  |  |  |
| --- | --- | --- | --- | --- | --- | --- | --- | --- | --- | --- | --- |
| | Estimate<br>(SE) | $p_{\text{FDR}}$ (raw<br>$p$-value) | Estimate | $p$-<br>value | Intercept<br>$p$-value | Raw<br>estimate | Raw $p$-<br>value | Outlier<br>Estimate | Outlier $p$-<br>value | Global $p$-<br>value | Distortion<br>$p$-value |
| BMI | -0.006<br>(0.054) | 1.0 (0.92) | 0.043 (0.10) | 0.67 | 0.57 | -0.006<br>(0.054) | 0.92 | -0.016<br>(0.043) | 0.73 | **0.012** | 0.24 |
| WHR | 0.018<br>(0.007) | 0.078<br>(0.013) | 0.023<br>(0.014) | 0.10 | 0.67 | 0.018 | 0.041 | - | - | 0.075 | - |
| Glucose | 0.019<br>(0.018) | 1.0 (0.30) | 0.010<br>(0.034) | 0.77 | 0.76 | 0.019 | 0.34 | - | - | 0.29 | - |
| Diabetes | 0.004<br>(0.007) | 1.0 (0.52) | -0.003<br>(0.012) | 0.80 | 0.46 | 0.004<br>(0.006) | 0.47 | - | - | 0.63 | - |
| HDL | -0.005<br>(0.004) | 1.0 (0.23) | -0.010<br>(0.007) | 0.15 | 0.36 | -0.005<br>(0.004) | 0.27 | -0.008<br>(0.005) | 0.17 | **0.021** | **<0.0002** |
| BMD | 0.008<br>(0.009) | 1.0 (0.41) | -0.005<br>(0.017) | 0.77 | 0.38 | 0.008<br>(0.009) | 0.44 | 0.003<br>(0.003) | 0.42 | 0.010 | 0.87 |

A significant finding of pleiotropy is indicated by the intercept $p$ -value in MR-Egger and the Global $p$ -value in MR-PRESSO. For MR-PRESSO the outlier test is only run if horizontal pleiotropy is observed. The distortion $p$ -value represents whether the outlier removal significantly reduces the horizontal pleiotropy.

### Reverse Mendelian randomisation analysis identifies BMI as a causal factor for GDF15 plasma levels

We applied reverse two-sample MR (as described above) using GDF15 plasma levels as the outcome variable to assess associations with BMI, WHR, glucose, diabetes, HDL cholesterol and BMD. GDF15 GWAS summary statistics were taken from the conditioned meta-analysis of GDF15 levels in FINRISK and INTERVAL. LD clumping was used to identify the genetic instruments (see Methods). We found a significant association between higher genetically-predicted BMI and higher GDF15 plasma levels (IVW estimate=0.089, *p*_FDR_=0.0072) but not any other tested trait (Table 4). Sensitivity analyses in MR-PRESSO but not MR-Egger analysis confirmed the association with BMI (*p*-value>0.05). MR-Egger identified horizontal pleiotropy with WHR, however, MR-PRESSO identified horizontal pleiotropy with every trait other than BMD. The finding of horizontal pleiotropy in MR-PRESSO but not MR-Egger suggests that the causal of an apparent causal effect of BMI on GDF15 plasma levels warrants replication in larger cohorts.

**Table 4.**
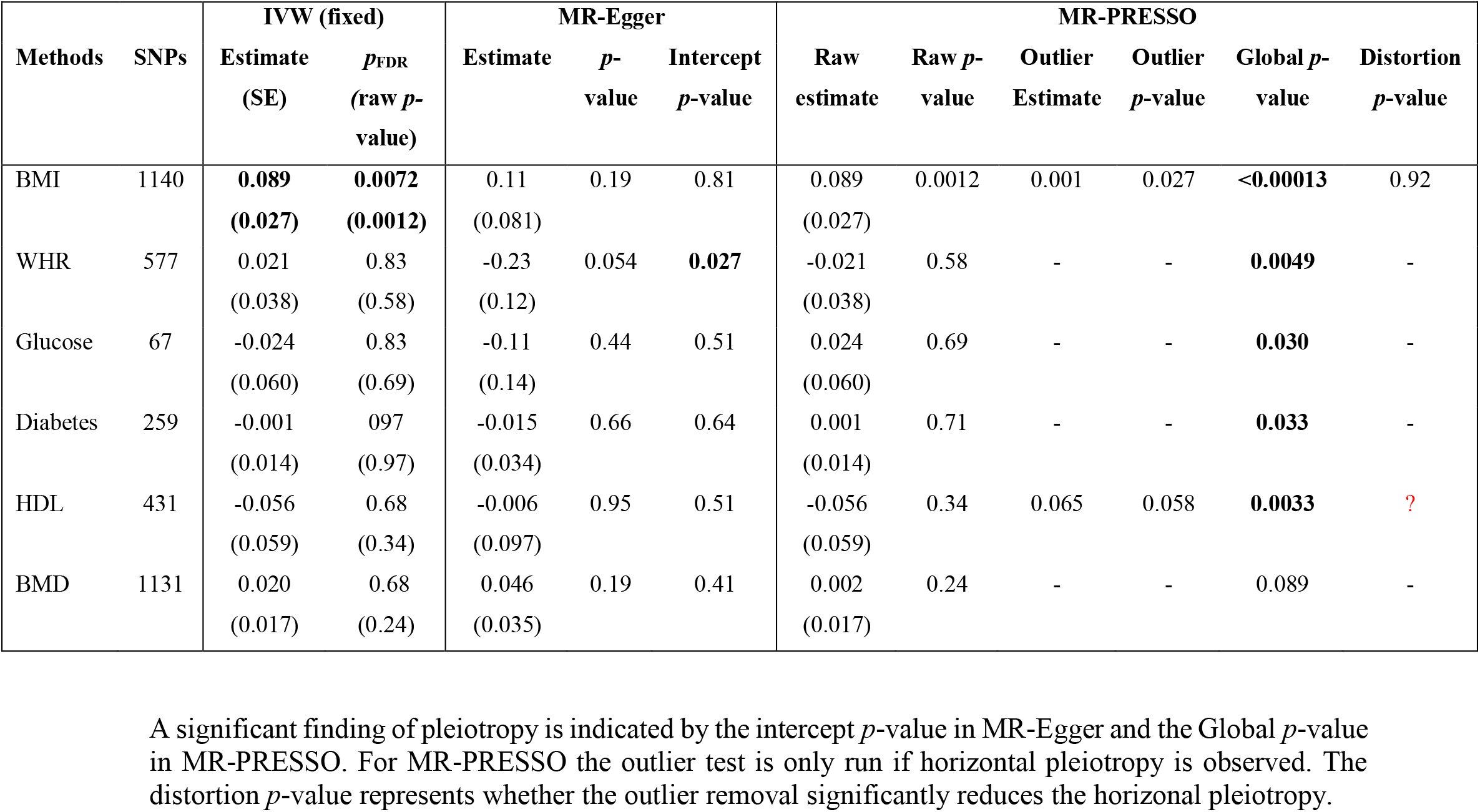
Reverse Mendelian randomization with GDF15 plasma levels as outcome.

| Methods | SNPs | IVW (fixed) |  | MR-Egger |  |  | MR-PRESSO |  |  |  |  |  |
| --- | --- | --- | --- | --- | --- | --- | --- | --- | --- | --- | --- | --- |
| | | Estimate<br>(SE) | $p_{\text{FDR}}$<br>(raw $p$-value) | Estimate | $p$-value | Intercept<br>$p$-value | Raw estimate | Raw $p$-value | Outlier Estimate | Outlier $p$-value | Global $p$-value | Distortion<br>$p$-value |
| BMI | 1140 | **0.089**<br>**(0.027)** | **0.0072**<br>**(0.0012)** | 0.11<br>(0.081) | 0.19 | 0.81 | 0.089<br>(0.027) | 0.0012 | 0.001 | 0.027 | **<0.00013** | 0.92 |
| WHR | 577 | 0.021<br>(0.038) | 0.83<br>(0.58) | -0.23<br>(0.12) | 0.054 | **0.027** | -0.021<br>(0.038) | 0.58 | - | - | **0.0049** | - |
| Glucose | 67 | -0.024<br>(0.060) | 0.83<br>(0.69) | -0.11<br>(0.14) | 0.44 | 0.51 | 0.024<br>(0.060) | 0.69 | - | - | **0.030** | - |
| Diabetes | 259 | -0.001<br>(0.014) | 0.97<br>(0.97) | -0.015<br>(0.034) | 0.66 | 0.64 | 0.001<br>(0.014) | 0.71 | - | - | **0.033** | - |
| HDL | 431 | -0.056<br>(0.059) | 0.68<br>(0.34) | -0.006<br>(0.097) | 0.95 | 0.51 | -0.056<br>(0.059) | 0.34 | 0.065 | 0.058 | **0.0033** | **?** |
| BMD | 1131 | 0.020<br>(0.017) | 0.68<br>(0.24) | 0.046<br>(0.035) | 0.19 | 0.41 | 0.002<br>(0.017) | 0.24 | - | - | 0.089 | - |

A significant finding of pleiotropy is indicated by the intercept $p$ -value in MR-Egger and the Global $p$ -value in MR-PRESSO. For MR-PRESSO the outlier test is only run if horizontal pleiotropy is observed. The distortion $p$ -value represents whether the outlier removal significantly reduces the horizontal pleiotropy.

### Effect of GDF15 protein-truncating variants on cardiometabolic traits in 302,388 participants of UK Biobank

Carriers of *GDF15* protein-truncating variants (PTVs) present an opportunity to explore the phenotypic consequences of predicted loss-of-function (LOF) of GDF15 on human disease. We analysed whole-exome sequencing data from 302,388 participants from the UK Biobank, of whom 109 carried *GDF15* PTVs in the heterozygous state (Supplementary Table S19). We assessed differences in BMI, WHR, glucose, BMD, HDL cholesterol and type 2 diabetes between carriers and non-carriers using the Mann-Whitney U test. The analysis was restricted to unrelated individuals of European ancestry, leaving 91 carriers of *GDF15* PTVs (male N=42, female N=49, mean age=56.3) and 40,000 randomly selected European-ancestry non-carriers (male N=20,000, female N=20,000, mean age=56.9). In line with the MR analyses we observed no differences in BMI (mean difference=0.1, *p-*value=0.90), WHR (male: mean difference=0.01, *p-*value=0.64, female: mean difference=0.01, *p-*value=0.55), glucose (mean difference=0.14, *p-*value=0.32), BMD (mean difference=0.02, *p-*value=0.82), HDL cholesterol (mean difference=0.08, *p-*value=0.08) or self-reported diabetes (N of diabetes in cases=2, *p-*value=0.12), suggesting that mono-allelic *GDF15* LOF does not have a strong effect on these traits.

## Discussion

In this study, we present a systematic phenotypic and genotypic analysis of GDF15 with a wide range of health outcomes and biomarkers. In line with previous findings, our analysis confirmed strong correlations of GDF15 plasma levels with a range of clinical parameters (e.g. age, smoking and BMI) and human diseases (e.g. diabetes, cardiovascular and respiratory disease), as well as several inflammatory biomarkers. As preclinical data in mice strongly implicated Gdf15 in the aetiology of obesity and glucose tolerance, we specifically investigated whether human genetic evidence supports these findings. Using data from large biobanks, we found that neither MR nor *GDF15* PTV analyses supported a causal role for GDF15 plasma levels within the normal range in influencing BMI, WHR, glucose, diabetes, HDL cholesterol or BMD in humans. Instead, we found that higher BMI may cause increases in GDF15 plasma levels highlighting the role of GDF15 as a likely marker of metabolic stress in humans.

Access to the FINRISK cohort provided a valuable opportunity to explore GDF15 plasma level associations with multiple phenotypes within a single large population. We found the strongest associations with all-cause mortality and cardiometabolic diseases (cardiovascular disease and diabetes), as well as cardiometabolic risk factors (e.g. hypertension, triglycerides, BMI), as previously reported (Bonaca et al., 2011; Brown et al., 2002; Daniels, Clopton, Laughlin, Maisel, & Barrett-Connor, 2011; Ho et al., 2012; Kempf et al., 2007; Khan et al., 2009; Lind et al., 2009; Rohatgi et al., 2012; Wiklund et al., 2010; Wollert et al., 2007). Our analyses also found that GDF15 plasma levels are is a strong predictor of incident diabetes and an independent predictor of all-cause mortality, cardiovascular disease and diabetes morbidity, suggesting its potential use as a pre-diabetic prognostic biomarker. We found associations with neoplasms of the lung and digestive system but not with other types of cancer. Our data further support the association of GDF15 plasma levels with inflammatory phenotypes and biomarkers (CRP, mid-regional pro-adrenomedullin) (Brown et al., 2007) and uncovered less well-described associations with respiratory disease and psychiatric disorders. Despite previous observations suggesting a role for GDF15 in anorexia (Borner, Shaulson, et al., 2020), we did not identify any association of GDF15 plasma levels with this trait. However, we note that statistical power may have been limited due to the low number of cases (Supplementary Table S5). To identify phenotype associations in an unbiased way, we performed analyses encompassing all phenotypes available applying covariates uniformly across all analyses. We note that covariates specific to a particular disease might have been omitted introducing bias or leading to inflation or deflation of statistics. We detected a strong association between GDF15 levels and smoking, which was recorded and adjusted in our analyses as a binary phenotype. Smoking is an important contributing factor in multiple diseases and it is therefore likely that adjustment as a continuous variable would have been able to better explain the contribution of smoking to the phenotypic association signals, especially in respiratory disease, lung cancer and some psychiatric disorders. These findings demonstrate that GDF15 plasma levels are a general marker for risk in multiple diseases and are associated with non-specific biomarkers such as CRP.

To define the genetic architecture of plasma GDF15 levels, we performed a GWAS meta-analysis in two large cohorts, FINRISK and INTERVAL, across three different assay platforms. A striking finding of this analysis was the substantial heterogeneity between variants across these studies, consistent with previous reports (Jiang et al., 2018). Exploring the LD between the variants displaying heterogeneity (Supplementary Figure S6) identified that a large proportion resided within an LD block encompassing a common missense variant, rs1058587 (p.H202D). This variant has been previously associated with hyperemesis gravidarum (Fejzo, Sazonova, et al., 2018) but the presence of this missense variant within this locus raises the possibility of epitope artefacts, indeed a previous study has identified epitope effects in this region (Fairlie et al., 2001). Therefore, it is highly likely that the inconsistency in GWAS results across this locus is driven by differences in the properties of the GDF15 assay used in each study (immunoluminometric assay in FINRISK, SomaScan and Olink assays in INTERVAL). Therefore, it may be beneficial to adopt a more consistent method for measuring GDF15 levels within the research environment. Approaches to assay protein levels that don’t involve binding to epitopes, such as mass spectrometry, will be informative for avoiding potential artefacts due to protein-altering variants in the future.

Our analyses did not reveal a causal role for GDF15 plasma levels with any of these cardiometabolic phenotypes. These results were further supported by the lack of association between these phenotypes and *GDF15* PTV carriers in the UK Biobank dataset. This is consistent with a recent large-scale, pan-ancestry exome-sequencing study of over 640,000 individuals exploring the associations of rare coding variants with BMI that did not report an association with GDF15 (Akbari et al., 2021). However, a meta-analysis completed by Cheung *et al*. (Ho et al., 2012) did identify significant associations of genetically-predicted GDF15 levels with HDL cholesterol (beta=-0.048, *p*-value=0.001) and estimated BMD (beta=0.026, *p*-value<0.001) which we did not replicate. The inclusion of differing covariates could have influenced the results of these analyses; our genetic analysis included only age, gender and principal components as covariates whereas the meta-analysis by Cheung *et al*. included age, gender, systolic blood pressure, antihypertensive medication use, diabetes mellitus and smoking status. Nevertheless, the larger sample size of the study presented here (n=14,099) offered considerably improved statistical power over this previous study (n=3,889).

This lack of causal association with GDF15 plasma levels suggests that GDF15 may behave differently in humans compared to rodents and non-human primates, where there are robust data demonstrating that GDF15 potently reduce body weight. GDF15 levels have also been demonstrated to provoke nausea-related pica and CTA responses in rat and mice, respectively (Borner, Shaulson, et al., 2020; Patel et al., 2019), as well as vomiting in shrews, *suncus murinus*, but studies in non-human primates report no signs of nausea (Xiong et al., 2017). Several clinical trials testing the safety and efficacy of GDF15 therapy have been planned or initiated but no data have yet been disclosed. Nevertheless, body weight loss in pregnant women suffering from hyperemesis gravidarium has been linked with higher plasma GDF15 levels (Fejzo, Arzy, et al., 2018). In FINRISK, information on nausea was not available but diseases with nausea as the main symptom were available (such as gastroesophageal reflux disease) and no significant association with GDF15 plasma levels was found (OR=1.1, p=0.63). In pregnancy GDF15 levels are found at much higher serum levels than normal (A. G. Moore et al., 2000) and it is therefore possible that at higher levels GDF15 may induce nausea. Further studies into the effect of higher-concentration GDF15 in humans and the outcomes of clinical trials will elucidate its role further.

As our analyses did not support a causal role for GDF15 plasma levels in the phenotypes examined, we sought to investigate if genetics supported the role for GDF15 as a consequence of these conditions. We conducted reverse MR using GDF15 plasma levels as the outcome and the previously assessed cardiometabolic traits and diseases as the exposures. We identified a significant causal association of BMI on GDF15 plasma levels, suggesting that higher GDF15 levels could be a consequence of higher BMI, supporting the recently reported role of GDF15 in response to stress (Patel et al., 2019). However, the contradictory findings of horizontal pleiotropy between MR-Egger and MR-PRESSO lead to residual uncertainty in the interpretation of the MR analysis. It is possible that the heterogeneity found in our study caused by the use of different assays is driving this pleiotropic finding. A recent report in a MR study of GDF15 levels performed by the SCALLOP consortium also found a significant causal effect of BMI (IVW effect=0.20, *p*-value=1×10^−7^) (Folkersen et al., 2020). Our replication of this finding strengthens the evidence that GDF15 levels are likely a consequence of differences in BMI rather than a cause. Further research will be required to determine conclusively the role of horizontal pleiotropy in this relationship.

Using valid genetic instruments in MR is of key importance as an invalid instrument would violate the assumptions of the model and lead to unreliable results. To minimise bias from differential assay binding, we conditioned on rs1058587 in our analyses, which could be overcautious. The association of rs1058587 with adiposity traits and hyperemesis gravidarum raises the possibility that GDF15 exerts a causal effect that is not mediated by altered plasma levels, such as via altered proteoforms and/or altered GFRAL binding. With the use of a wide range of GDF15 assays and the possibility that this missense variant is impacting the assay functionality in different assays to varying degrees, the generation of functional data quantifying assay performance is essential for understanding the contribution of this association to GDF15 plasma levels and improving the power of MR analyses. Similarly, as our *GDF15* PTV data is based solely on heterozygous carriers, it is possible that the other allele is compensating for the loss and further assessment of the impact of heterozygote *GDF15* PTVs on the presence of the protein will be required to validate our findings.

Taken together, our study provides a systematic and broad investigation of GDF15 phenotypic and genotypic associations, identifying possible epitope artefacts that could affect the data validity of GDF15 assays and introduce systematic bias in genetic analyses. Taking into account these biases, our genetic analyses did not support a causal association between GDF15 plasma levels and obesity and diabetes in humans. Conversely, we found that increased GDF15 plasma levels may be a consequence of higher BMI. We suggest the strong and wide-ranging associations between GDF15 plasma levels and health outcomes observed predominantly represent secondary effects of the underlying disease process, implicating GDF15 as a stress-induced biomarker. The notion that GDF15 may be merely a predictive marker rather than a causal factor for metabolic diseases draws parallels to CRP in cardiovascular disease. Here, epidemiological studies revealed that elevated plasma CRP levels are consistently associated with increased risks of atherosclerosis and ischemic vascular disease, but Mendelian randomization studies do not show a causal relationship (Davey Smith et al., 2005). Our results do not support GDF15 plasma levels as a causal factor at normal human plasma levels for obesity or its related cardiometabolic diseases.

## Materials and Methods

### Study population and phenotypes

#### FINRISK

This study was carried out in accordance with the recommendations of the Declaration of Helsinki. All participants of studies have given written informed consent. FINRISK study was approved by the Ethics Committee of Helsinki and Uusimaa Hospital District.

The FINRISK cohort comprises a cross-sectional population survey carried out over a 40-year period from the year 1972 across multiple regions in Finland. The study aimed to assess the risk factors of chronic diseases and health behaviours in a working age population (Borodulin et al., 2018; Vartiainen et al., 2000) and consisted of 6,000-8,800 individuals per survey. Measurement of GDF15 plasma levels using an immunoluminometric assay was included for participants from the 1997 recruitment cohort. Participants were additionally matched to their electronic health records giving access to longitudinal prescription records, death records and diagnosis history. The cohort characteristics are summarized in Supplementary Table S1. In total, 6,610 Finnish individuals from the FINRISK 1997 cohort with available GDF15 plasma concentrations and up to 676 disease outcomes were included in this analysis.

Nurse assessment/interview, self-report data and blood samples were all collected at various time points during the study (see Supplementary Materials). A group of clinicians working in the FinnGen consortium constructed 676 disease endpoints combining information from multiple health registries (T et al., 2020). Both ICD8, ICD9 and ICD10 codes were utilized in the disease endpoint definitions. Genotyping was carried out for in 6,538 individuals who were recruited in 1997, for more information see Suplementary Materials.

#### INTERVAL

The INTERVAL study is a prospective cohort study comprising approximately 50,000 participants nested within a pragmatic randomized trial of blood donors [33]. Between 2012 and 2014, blood donors aged 18 years and older were recruited at 25 NHSBT (National Health Service Blood and Transplant) donor centres across England (see Supplementary Materials). Informed consent was obtained from all participants and the INTERVAL study was approved by the National Research Ethics Service (11/EE/0538).

For the SOMAscan assays, two non-overlapping subcohorts of 2,731 and 831 participants were randomly selected, of which 3,301 participants (2,481 and 820 in the two subcohorts) remained after genetic quality control.

For the Olink assay, protein measurements were conducted in an additional subcohort of 4,998 INTERVAL participants aged over 50 at baseline. 4,987 samples passed quality control for this panel and were included in the analyses.

For information on genotyping in the INTERVAl cohort see Suplementary Materials

#### UK Biobank

UK Biobank is a population-based cohort consisting of ∼500,000 individuals with participants recruited between 2006-2010 and aged between 40-69. Recruitment and cohort information has been previously described (Sudlow et al., 2015). Electronic health records, self-report questionnaires, diet and lifestyle information and biomarker data is available on this cohort. Here, we utilised body mass index (BMI) and waist-hip-ratio (WHR), which were measured during a nurse assessment, as well as diabetes information. Touchscreen questionnaire diabetic patients were applied in this study rather than ICD10 diagnosed in order to increase patient numbers, given the small subgroup of individuals being examined. Details on the exome sequencing are available in the Supplementary Materials.

### Laboratory methods for GDF15 measurement

#### FINRISK

Blood samples were collected after an advisory 4-hour fast, immediately centrifuged and then stored at -70°C until GDF15 measurement which was undertaken using an immunoluminometric assay (ILMA) with a limit of detection of 24 ng/L and a linear range from 200 to 50.000 ng/L (Kempf et al., 2007). The ILMA is technically identical to immunoradiometric assay (IRMA) (Horn, Geldszus, Potter, von zur Muhlen, & Brabant, 1996) except that the GDF15 detection antibody was labelled with acridinium ester and assay results were quantified in a luminometer (Berthold). ILMA and IRMA use an antibody that binds over a sequence of Ala197-Ile308. GDF-15 concentrations measured with the ILMA and IRMA are very similar [(ILMA concentration/ILMA concentration) × 100% = 97.8 ± 1.3%] and closely correlated (*r* 2 = 0.99. *p* < 0.001. *n* = 31 samples) (H). Head-to-head comparison of IRMA with the clinical ‘gold standard’ Roche assay have been previously conducted and reported GDF15 levels were comparable (Wollert et al., 2017).

#### INTERVAL

Blood sample collection procedures have been described previously (C. Moore et al., 2014). Blood samples were collected in 6-ml EDTA tubes and transferred at ambient temperature to UK Biocentre (Stockport, UK) for processing. Plasma was extracted by centrifugation into two 0.8-ml plasma aliquots and stored at -80°C before use.

The procedures for obtaining protein measurements using the SOMAscan assay have also been described previously (Sun et al., 2018). Briefly, SOMAscan is a multiplexed, aptamer-based approach that was used to measure the relative concentrations of 3,622 plasma proteins or protein complexes using modified aptamers (‘SOMAmer reagents’, hereafter referred to as SOMAmers). Aliquots of 150 µl of plasma from INTERVAL baseline samples were sent on dry ice to SomaLogic Inc. (Boulder, Colorado, US) for protein measurement. Modified single-stranded DNA SOMAmers were used to bind to specific protein targets and these are then quantified using a DNA microarray. Protein concentrations are quantified as relative fluorescent units. Quality control (QC) was completed at the sample and SOMAmer levels.

The Olink assay uses pairs of monoclonal antibodies (or a single polyclonal antibody) to bind each protein target, and then uses proximity extension followed by qPCR to quantify protein abundance. Aliquots of plasma from samples taken from INTERVAL participants at the 2-year follow-up survey were shipped on dry ice to Olink Proteomics (Uppsala, Sweden) for assay. Protein concentrations were recorded as normalised relative protein abundances (‘NPX’).

### Genome-wide association analysis

#### FINRISK

For the FINRISK cohort genome-wide association analyses were performed for 5817 individuals with plasma GDF15 concentrations available. A normal distribution of GDF15 plasma concentrations was achieved through applying an inverse normal transformation. Multidimensional scaling was done for genetic data of both studies using PLINK version 1.07 (Purcell et al., 2007). Only good quality autosomal markers passing the following criteria: imputation informativeness >0.7, and MAF>0.001, were included in further evaluations. Frequentist test with method ‘expected’, assuming an additive genetic model, was performed using SNPTEST v2 (Marchini, Howie, Myers, McVean, & Donnelly, 2007). Results were adjusted for the first five principal components of the genetic data to account for the population stratification and for gender, age and genotyping set. The Quantile-Quantile and Manhattan plots and regional plots were created using R-2.11 to visualize genome-wide association results. The genomic positions indicated throughout this study are based on NCBI human genome build 37.

#### INTERVAL

The GWAS for INTERVAL-SOMAscan has previously been described in detail (Sun et al., 2018). Relative protein abundances were first natural log-transformed within each subcohort. Log-transformed protein levels were then adjusted in a linear regression for age, sex, duration between blood draw and processing (binary, ≤1 day/>1day) and the first three principal components of ancestry from multi-dimensional scaling. A normal distribution of the protein residuals from this linear regression was achieved through rank-inverse normal transformation and these were used as phenotypes for association testing. Simple linear regression assuming an additive genetic model was used to test for genetic associations. Genotype uncertainty was accounted for by carrying out association tests on allelic dosages (‘method expected’ option) using SNPTEST v2.5.2 [51].

For the INTERVAL-Olink GWAS, normalized protein levels (‘NPX’) were first regressed on age, sex, plate, time from blood draw to processing (in days), and season (categorical: ‘Spring’, ‘Summer’, ‘Autumn’, ‘Winter’). The residuals from this linear regression were rank-inverse normalized. The rank-inverse normalized residuals were then fitted in a linear regression model adjusted for ancestry by including the first three components of multi-dimensional scaling as covariates. GWAS was conducted using SNPTEST v2.5.2.

### Fine mapping

Fine mapping was performed using FINEMAP program (Benner et al., 2016). FINEMAP can potentially identify sets of variants with more evidence of being causal than those highlighted by a stepwise conditional analysis. The FINEMAP provides (1) a list of potential causal configurations together with their posterior probabilities and Bayes factors and, (2) for each variant, the posterior probability and Bayes factor of being causal. FINEMAP was applied with its default settings. LD between SNPs for the conditional analysis was combined using the formula: (N1 * LD1 + N2 * LD2 + N3 * LD3) / (N1 + N2 + N3), where N represents the number of individuals in the GWAS in each cohort, LD represents the R^2^ value and 1, 2 and 3 represent the three different cohorts: FINRISK, INTERVAL-SomaScan and INTERVAL-Olink.

### Meta-analysis

Meta-analysis was completed on the chromosome 19 locus around the GDF15 gene in METAL (Willer, Li, & Abecasis, 2010) using the Inverse-variance Weighted method. A total of 3,799 SNPs were found in common across FINRISK, INTERVAL-SomaScan and INTERVAL-Olink.

### Statistical analysis

#### Association study between GDF15 levels and disease endpoints and quantitative biomarkers

To examine the association of GDF15 with disease endpoints and quantitative biomarkers, logistic and linear regression models were used, respectively. Inverse-variance transformed GDF15 levels were used in the analyses due to right-skewed distribution. Association of GDF15 with disease endpoints and quantitative biomarkers were examined in 1) age- and sex 2) age, sex and BMI 3) age, sex and smoking -adjusted models. Analyses using linear- and logistic regression models were performed using R2.11 (see URLs). False discovery rate (FDR) was applied to test for multiple test correction.

#### Survival analysis

All analyses were performed in R version 3.4.0. Cox proportional hazard regression model was conducted to identify predictors of outcomes during 10 years. The survival curves for a Cox proportional hazards model were used to illustrate the timing of the death, type 2 diabetes and CVD events during 10 years in relation to GDF15 quartiles and statistical assessment between upper quartile and other quartiles was preformed using cox proportional hazard regression model. To inspect the validity of the cox-model, the test of the proportional hazards assumption for a Cox regression model was used. Hosmer-Lemeshow Goodness of Fit (GOF) test was used to determine whether models were calibrated with similar expected and observed event rates in both low- and high-risk individuals.

#### Wilcoxon test for prevalent and incident diabetes

Wilcoxon rank sum test (Mann–Whitney U test) was performed, in R using function “wilcox.test” from stats package, to test whether the distributions of incident type 2 diabetes case group, prevalent type 2 diabetes case group and control group were systematically different from one another.

#### Mendelian Randomisation

We applied MR (Bowden et al., 2015) to explore causality of GDF15 with BMI, WHR, glucose, type 2 diabetes, HDL cholesterol and BMD. We additionally explored these relationship in reverse; with GDF15 as an outcome and other traits as the exposure. GWAS summary statistics for GDF15 were taken from the conditional meta-analysis of FINRISK and INTERVAL for the forward MR and from FINRISK only for the reverse MR. GWAS in INTERVAL being completed on chromosome 19 only whereas GWAS in FINRISK were completed on all chromosomes. Summary statistics for the other traits were obtained from large public data resources: BMI (n=590,827) (Yengo L et al., 2018), WHR (n=485,486) (Shungin et al., 2015), glucose (n=314,916) (Neale, 2018), type 2 diabetes (n cases=74,124, n controls=824,006) (Mahajan et al., 2018), HDL cholesterol (n=315,133) (Neale, 2018) and BMD (n=426,824) (Morris et al., 2019). For BMI, MR with GDF15 as the exposure was applied using summary statistics from UK Biobank only (n=361,194) as the LD clumped variants were not available in the larger meta-analysis, that was utilised for reverse MR. We utilised LD clumping to identify independent genetic variants in summary statistics and applied these as genetic instruments in MR analyses. MR analysis was run in R (version 4.0.2) using the package ‘MendelianRandomization’. The inverse variance weighted method (IVW) from this package was applied to test for causality and a sensitivity analysis using MR-Egger to testing for horizontal pleiotropy. A finding of significant causality is indicated with the IVW *p*-value and a significant MR-Egger intercept *p*-value indicates a finding of horizontal pleiotropy. MR-PRESSO (Mendelian randomization pleiotropy residual sum and outlier) (Verbanck et al., 2018) was additionally applied as a further sensitivity analysis to test for horizontal pleiotropy and was run in R using the ‘MRPRESSO’ package. The Global *p*-value indicates a significant finding of horizontal pleiotropy which instigates removal of outliers from the analysis. MR-PRESSO then re-runs IVW testing and gives a significant distortion *p*-value to indicate if the removal of outliers reduced the horizontal pleiotropy found in the analysis.

### GDF15 PTV analysis

#### Exome data

Individuals with *GDF15* PTVs were identified by extracting variants annotated as protein truncating variants (including exon loss variants, frameshift variants, start lost, stop gained, stop lost, splice acceptor variants, splice donor variant, rare amino acid variant, transcript ablation, gene fusion and bidirectional gene fusion). Supplementary Table S19 demonstrates the frequency of LOF variants in each ancestry. From the individuals that did not carry *GDF15* PTV variants 20,000 males and 20,000 females were randomly extracted as controls.

#### Mann Whitney U test

Analysis was completed in R (version 3.2.4) using “wilcox.test” and models compared BMI, WHR (in males and females separately), diabetic status (self-reported), BMD and HDL in *GDF15* PTV carriers and non-carriers.

## Data Availability

Summary data produced in the present study are available upon reasonable request to the authors

## Acknowledgements

We thank the participants and investigators in the UK Biobank study who made this work possible (Resource Application Number 26041). We thank the UK Biobank Exome Sequencing Consortium (UKB-ESC) members AbbVie, Alnylam Pharmaceuticals, AstraZeneca, Biogen, Bristol-Myers Squibb, Pfizer, Regeneron and Takeda for funding the generation of the data and Regeneron Genetics Center for completing the sequencing and initial quality control of the exome sequencing data. We thank the AstraZeneca Centre for Genomics Research Analytics and Informatics team for processing and analysis of sequencing data.

Participants in the INTERVAL randomised controlled trial were recruited with the active collaboration of NHS Blood and Transplant England (www.nhsbt.nhs.uk), which has supported field work and other elements of the trial. DNA extraction and genotyping was co-funded by the National Institute for Health Research (NIHR), the NIHR BioResource (http://bioresource.nihr.ac.uk) and the NIHR Cambridge Biomedical Research Centre (BRC-1215-20014) [*]. SomaLogic assays were funded by Merck and the NIHR Cambridge BRC (BRC-1215-20014) [*]. The academic coordinating centre for INTERVAL was supported by core funding from: NIHR Blood and Transplant Research Unit in Donor Health and Genomics (NIHR BTRU-2014-10024), UK Medical Research Council (MR/L003120/1), British Heart Foundation (SP/09/002; RG/13/13/30194; RG/18/13/33946) and the NIHR Cambridge BRC (BRC-1215-20014) [*]. A complete list of the investigators and contributors to the INTERVAL trial is provided in reference [**]. The academic coordinating centre would like to thank blood donor centre staff and blood donors for participating in the INTERVAL trial.

This work was supported by Health Data Research UK, which is funded by the UK Medical Research Council, Engineering and Physical Sciences Research Council, Economic and Social Research Council, Department of Health and Social Care (England), Chief Scientist Office of the Scottish Government Health and Social Care Directorates, Health and Social Care Research and Development Division (Welsh Government), Public Health Agency (Northern Ireland), British Heart Foundation and Wellcome.

This work was conducted as part of an alliance between the University of Cambridge and the AstraZeneca Centre for Genomics Research (AZ Ref: 10033507).

The Genotype-Tissue Expression (GTEx) Project was supported by the Common Fund of the Office of the Director of the National Institutes of Health, and by NCI, NHGRI, NHLBI, NIDA, NIMH, and NINDS. The data used for the analyses described in this manuscript were obtained from the GTEx Portal on 08/12/2021.

*The views expressed are those of the author(s) and not necessarily those of the NIHR or the Department of Health and Social Care.

**Di Angelantonio E, Thompson SG, Kaptoge SK, Moore C, Walker M, Armitage J, Ouwehand WH, Roberts DJ, Danesh J, INTERVAL Trial Group. Efficiency and safety of varying the frequency of whole blood donation (INTERVAL): a randomised trial of 45 000 donors. Lancet. 2017 Nov 25;390(10110):2360-2371.

## Author contributions

## REFERENCES

Adela, R., & Banerjee, S. K. (2015). GDF-15 as a Target and Biomarker for Diabetes and Cardiovascular Diseases: A Translational Prospective. J Diabetes Res, 2015, 490842. doi:10.1155/2015/490842

Akbari, P., Gilani, A., Sosina, O., Kosmicki, J. A., Khrimian, L., Fang, Y. Y., … Lotta, L. A. (2021). Sequencing of 640,000 exomes identifies GPR75 variants associated with protection from obesity. Science, 373(6550). doi:10.1126/science.abf8683

Bonaca, M. P., Morrow, D. A., Braunwald, E., Cannon, C. P., Jiang, S., Breher, S., … Wollert, K. C. (2011). Growth differentiation factor-15 and risk of recurrent events in patients stabilized after acute coronary syndrome: observations from PROVE IT-TIMI 22. Arterioscler Thromb Vasc Biol, 31(1), 203–210. doi:10.1161/ATVBAHA.110.213512

Bootcov, M. R., Bauskin, A. R., Valenzuela, S. M., Moore, A. G., Bansal, M., He, X. Y., … Breit, S. N. (1997). MIC-1, a novel macrophage inhibitory cytokine, is a divergent member of the TGF-beta superfamily. Proc Natl Acad Sci U S A, 94(21), 11514–11519. doi:10.1073/pnas.94.21.11514

Borner, T., Shaulson, E. D., Ghidewon, M. Y., Barnett, A. B., Horn, C. C., Doyle, R. P., … De Jonghe, B. C. (2020). GDF15 Induces Anorexia through Nausea and Emesis. Cell Metab, 31(2), 351–362 e355. doi:10.1016/j.cmet.2019.12.004

Borner, T., Wald, H. S., Ghidewon, M. Y., Zhang, B., Wu, Z., De Jonghe, B. C., … Grill, H. J. (2020). GDF15 Induces an Aversive Visceral Malaise State that Drives Anorexia and Weight Loss. Cell Rep, 31(3), 107543. doi:10.1016/j.celrep.2020.107543

Borodulin, K., Tolonen, H., Jousilahti, P., Jula, A., Juolevi, A., Koskinen, S., … Vartiainen, E. (2018). Cohort Profile: The National FINRISK Study. Int J Epidemiol, 47(3), 696–696i. doi:10.1093/ije/dyx239

Bowden, J., Davey Smith, G., & Burgess, S. (2015). Mendelian randomization with invalid instruments: effect estimation and bias detection through Egger regression. Int J Epidemiol, 44(2), 512–525. doi:10.1093/ije/dyv080

Brown, D. A., Breit, S. N., Buring, J., Fairlie, W. D., Bauskin, A. R., Liu, T., & Ridker, P. M. (2002). Concentration in plasma of macrophage inhibitory cytokine-1 and risk of cardiovascular events in women: a nested case-control study. Lancet, 359(9324), 2159–2163. doi:10.1016/S0140-6736(02)09093-1

Brown, D. A., Moore, J., Johnen, H., Smeets, T. J., Bauskin, A. R., Kuffner, T., … Breit, S. N. (2007). Serum macrophage inhibitory cytokine 1 in rheumatoid arthritis: a potential marker of erosive joint destruction. Arthritis Rheum, 56(3), 753–764. doi:10.1002/art.22410

Cheung, C. L., Tan, K. C. B., Au, P. C. M., Li, G. H. Y., & Cheung, B. M. Y. (2019). Evaluation of GDF15 as a therapeutic target of cardiometabolic diseases in human: A Mendelian randomization study. EBioMedicine, 41, 85–90. doi:10.1016/j.ebiom.2019.02.021

Coll, A. P., Chen, M., Taskar, P., Rimmington, D., Patel, S., Tadross, J. A., … O’Rahilly, S. (2020). GDF15 mediates the effects of metformin on body weight and energy balance. Nature, 578(7795), 444–448. doi:10.1038/s41586-019-1911-y

Daniels, L. B., Clopton, P., Laughlin, G. A., Maisel, A. S., & Barrett-Connor, E. (2011). Growth-differentiation factor-15 is a robust, independent predictor of 11-year mortality risk in community-dwelling older adults: the Rancho Bernardo Study. Circulation, 123(19), 2101–2110. doi:10.1161/CIRCULATIONAHA.110.979740

Davey Smith, G., Lawlor, D. A., Harbord, R., Timpson, N., Rumley, A., Lowe, G. D., … Ebrahim, S. (2005). Association of C-reactive protein with blood pressure and hypertension: life course confounding and mendelian randomization tests of causality. Arterioscler Thromb Vasc Biol, 25(5), 1051–1056. doi:10.1161/01.ATV.0000160351.95181.d0

Day, E. A., Ford, R. J., Smith, B. K., Mohammadi-Shemirani, P., Morrow, M. R., Gutgesell, R. M., … Steinberg, G. R. (2019). Metformin-induced increases in GDF15 are important for suppressing appetite and promoting weight loss. Nat Metab, 1(12), 1202–1208. doi:10.1038/s42255-019-0146-4

Emmerson, P. J., Duffin, K. L., Chintharlapalli, S., & Wu, X. (2018). GDF15 and Growth Control. Front Physiol, 9, 1712. doi:10.3389/fphys.2018.01712

Emmerson, P. J., Wang, F., Du, Y., Liu, Q., Pickard, R. T., Gonciarz, M. D., … Wu, X. (2017). The metabolic effects of GDF15 are mediated by the orphan receptor GFRAL. Nat Med, 23(10), 1215–1219. doi:10.1038/nm.4393

Fairlie, W. D., Russell, P. K., Wu, W. M., Moore, A. G., Zhang, H. P., Brown, P. K., … Breit, S. N. (2001). Epitope mapping of the transforming growth factor-beta superfamily protein, macrophage inhibitory cytokine-1 (MIC-1): identification of at least five distinct epitope specificities. Biochemistry, 40(1), 65–73. doi:10.1021/bi001064p

Fejzo, M. S., Arzy, D., Tian, R., MacGibbon, K. W., & Mullin, P. M. (2018). Evidence GDF15 Plays a Role in Familial and Recurrent Hyperemesis Gravidarum. Geburtshilfe Frauenheilkd, 78(9), 866–870. doi:10.1055/a-0661-0287

Fejzo, M. S., Sazonova, O. V., Sathirapongsasuti, J. F., Hallgrimsdottir, I. B., Vacic, V., MacGibbon, K. W., … andMe Research, T. (2018). Placenta and appetite genes GDF15 and IGFBP7 are associated with hyperemesis gravidarum. Nat Commun, 9(1), 1178. doi:10.1038/s41467-018-03258-0

Folkersen, L., Gustafsson, S., Wang, Q., Hansen, D. H., Hedman, A. K., Schork, A., … Malarstig, A. (2020). Genomic and drug target evaluation of 90 cardiovascular proteins in 30,931 individuals. Nat Metab, 2(10), 1135–1148. doi:10.1038/s42255-020-00287-2

H, T. Dissertation.

Ho, J. E., Mahajan, A., Chen, M. H., Larson, M. G., McCabe, E. L., Ghorbani, A., … Wang, T. J. (2012). Clinical and genetic correlates of growth differentiation factor 15 in the community. Clin Chem, 58(11), 1582–1591. doi:10.1373/clinchem.2012.190322

Holmes, M. V., Lange, L. A., Palmer, T., Lanktree, M. B., North, K. E., Almoguera, B., … Keating, B. J. (2014). Causal effects of body mass index on cardiometabolic traits and events: a Mendelian randomization analysis. Am J Hum Genet, 94(2), 198–208. doi:10.1016/j.ajhg.2013.12.014

Horn, R., Geldszus, R., Potter, E., von zur Muhlen, A., & Brabant, G. (1996). Radioimmunoassay for the detection of leptin in human serum. Exp Clin Endocrinol Diabetes, 104(6), 454–458. doi:10.1055/s-0029-1211484

Hsu, J. Y., Crawley, S., Chen, M., Ayupova, D. A., Lindhout, D. A., Higbee, J., … Allan, B. B. (2017). Non-homeostatic body weight regulation through a brainstem-restricted receptor for GDF15. Nature, 550(7675), 255–259. doi:10.1038/nature24042

Jackson, C. E., Haig, C., Welsh, P., Dalzell, J. R., Tsorlalis, I. K., McConnachie, A., … McMurray, J. J. (2016). The incremental prognostic and clinical value of multiple novel biomarkers in heart failure. Eur J Heart Fail, 18(12), 1491–1498. doi:10.1002/ejhf.543

Jammah, A. A. (2015). Endocrine and metabolic complications after bariatric surgery. Saudi J Gastroenterol, 21(5), 269–277. doi:10.4103/1319-3767.164183

Jiang, J., Thalamuthu, A., Ho, J. E., Mahajan, A., Ek, W. E., Brown, D. A., … Mather, K. A. (2018). A Meta-Analysis of Genome-Wide Association Studies of Growth Differentiation Factor-15 Concentration in Blood. Front Genet, 9, 97. doi:10.3389/fgene.2018.00097

Johnen, H., Lin, S., Kuffner, T., Brown, D. A., Tsai, V. W., Bauskin, A. R., … Breit, S. N. (2007). Tumor-induced anorexia and weight loss are mediated by the TGF-beta superfamily cytokine MIC-1. Nat Med, 13(11), 1333–1340. doi:10.1038/nm1677

Kempf, T., Eden, M., Strelau, J., Naguib, M., Willenbockel, C., Tongers, J., … Wollert, K. C. (2006). The transforming growth factor-beta superfamily member growth-differentiation factor-15 protects the heart from ischemia/reperfusion injury. Circ Res, 98(3), 351–360. doi:10.1161/01.RES.0000202805.73038.48

Kempf, T., Horn-Wichmann, R., Brabant, G., Peter, T., Allhoff, T., Klein, G., … Wollert, K. C. (2007). Circulating concentrations of growth-differentiation factor 15 in apparently healthy elderly individuals and patients with chronic heart failure as assessed by a new immunoradiometric sandwich assay. Clin Chem, 53(2), 284–291. doi:10.1373/clinchem.2006.076828

Khan, S. Q., Ng, K., Dhillon, O., Kelly, D., Quinn, P., Squire, I. B., … Ng, L. L. (2009). Growth differentiation factor-15 as a prognostic marker in patients with acute myocardial infarction. Eur Heart J, 30(9), 1057–1065. doi:10.1093/eurheartj/ehn600

Lind, L., Wallentin, L., Kempf, T., Tapken, H., Quint, A., Lindahl, B., … Wollert, K. C. (2009). Growth-differentiation factor-15 is an independent marker of cardiovascular dysfunction and disease in the elderly: results from the Prospective Investigation of the Vasculature in Uppsala Seniors (PIVUS) Study. Eur Heart J, 30(19), 2346–2353. doi:10.1093/eurheartj/ehp261

Lockhart, S. M., Saudek, V., & O’Rahilly, S. (2020). GDF15: A hormone conveying somatic distress to the brain. Endocr Rev. doi:10.1210/endrev/bnaa007

Madonna, R., Cevik, C., Nasser, M., & De Caterina, R. (2012). Hepatocyte growth factor: molecular biomarker and player in cardioprotection and cardiovascular regeneration. Thromb Haemost, 107(4), 656–661. doi:10.1160/TH11-10-0711

Mahajan, A., Taliun, D., Thurner, M., Robertson, N. R., Torres, J. M., Rayner, N. W., … McCarthy, M. I. (2018). Fine-mapping type 2 diabetes loci to single-variant resolution using high-density imputation and islet-specific epigenome maps. Nat Genet, 50(11), 1505–1513. doi:10.1038/s41588-018-0241-6

Marchini, J., Howie, B., Myers, S., McVean, G., & Donnelly, P. (2007). A new multipoint method for genome-wide association studies by imputation of genotypes. Nat Genet, 39(7), 906–913. doi:10.1038/ng2088

Matsumoto, K., Umitsu, M., De Silva, D. M., Roy, A., & Bottaro, D. P. (2017). Hepatocyte growth factor/MET in cancer progression and biomarker discovery. Cancer Sci, 108(3), 296–307. doi:10.1111/cas.13156

Mesureur, L., & Arvanitakis, M. (2017). Metabolic and nutritional complications of bariatric surgery : a review. Acta Gastroenterol Belg, 80(4), 515–525.

Miller, V. M., Redfield, M. M., & McConnell, J. P. (2007). Use of BNP and CRP as biomarkers in assessing cardiovascular disease: diagnosis versus risk. Curr Vasc Pharmacol, 5(1), 15–25. doi:10.2174/157016107779317251

Moore, A. G., Brown, D. A., Fairlie, W. D., Bauskin, A. R., Brown, P. K., Munier, M. L., … Breit, S. N. (2000). The transforming growth factor-ss superfamily cytokine macrophage inhibitory cytokine-1 is present in high concentrations in the serum of pregnant women. J Clin Endocrinol Metab, 85(12), 4781–4788. doi:10.1210/jcem.85.12.7007

Moore, C., Sambrook, J., Walker, M., Tolkien, Z., Kaptoge, S., Allen, D., … Danesh, J. (2014). The INTERVAL trial to determine whether intervals between blood donations can be safely and acceptably decreased to optimise blood supply: study protocol for a randomised controlled trial. Trials, 15, 363. doi:10.1186/1745-6215-15-363

Morris, J. A., Kemp, J. P., Youlten, S. E., Laurent, L., Logan, J. G., Chai, R. C., … Richards, J. B. (2019). An atlas of genetic influences on osteoporosis in humans and mice. Nat Genet, 51(2), 258–266. doi:10.1038/s41588-018-0302-x

Mullican, S. E., Lin-Schmidt, X., Chin, C. N., Chavez, J. A., Furman, J. L., Armstrong, A. A., … Rangwala, S. M. (2017). GFRAL is the receptor for GDF15 and the ligand promotes weight loss in mice and nonhuman primates. Nat Med, 23(10), 1150–1157. doi:10.1038/nm.4392

Nair, V., Robinson-Cohen, C., Smith, M. R., Bellovich, K. A., Bhat, Z. Y., Bobadilla, M., … Bansal, N. (2017). Growth Differentiation Factor-15 and Risk of CKD Progression. J Am Soc Nephrol, 28(7), 2233–2240. doi:10.1681/ASN.2016080919

Neale, B. (2018). GWAS results.

O’Rahilly, S. (2017). GDF15-From Biomarker to Allostatic Hormone. Cell Metab, 26(6), 807–808. doi:10.1016/j.cmet.2017.10.017

Organisaton., W. H. Global Health Observatory (GBO) data: Obesity.

Patel, S., Alvarez-Guaita, A., Melvin, A., Rimmington, D., Dattilo, A., Miedzybrodzka, E. L., … O’Rahilly, S. (2019). GDF15 Provides an Endocrine Signal of Nutritional Stress in Mice and Humans. Cell Metab, 29(3), 707–718 e708. doi:10.1016/j.cmet.2018.12.016

Peacock, W. F. (2014). Novel biomarkers in acute heart failure: MR-pro-adrenomedullin. Clin Chem Lab Med, 52(10), 1433–1435. doi:10.1515/cclm-2014-0222

Pietzner, M., Wheeler, E., Carrasco-Zanini, J., Raffler, J., Kerrison, N. D., Oerton, E., … Langenberg, C. (2020). Genetic architecture of host proteins interacting with SARS-CoV-2. BioRxiv, doi: https://doi.org/10.1101/2020.1107.1101.182709doi.

Purcell, S., Neale, B., Todd-Brown, K., Thomas, L., Ferreira, M. A., Bender, D., … Sham, P. C. (2007). PLINK: a tool set for whole-genome association and population-based linkage analyses. Am J Hum Genet, 81(3), 559–575. doi:10.1086/519795

Rohatgi, A., Patel, P., Das, S. R., Ayers, C. R., Khera, A., Martinez-Rumayor, A., … de Lemos, J. A. (2012). Association of growth differentiation factor-15 with coronary atherosclerosis and mortality in a young, multiethnic population: observations from the Dallas Heart Study. Clin Chem, 58(1), 172–182. doi:10.1373/clinchem.2011.171926

Shungin, D., Winkler, T. W., Croteau-Chonka, D. C., Ferreira, T., Locke, A. E., Magi, R., … Mohlke, K. L. (2015). New genetic loci link adipose and insulin biology to body fat distribution. Nature, 518(7538), 187–196. doi:10.1038/nature14132

Sudlow, C., Gallacher, J., Allen, N., Beral, V., Burton, P., Danesh, J., … Collins, R. (2015). UK biobank: an open access resource for identifying the causes of a wide range of complex diseases of middle and old age. PLoS Med, 12(3), e1001779. doi:10.1371/journal.pmed.1001779

Sun, B. B., Maranville, J. C., Peters, J. E., Stacey, D., Staley, J. R., Blackshaw, J., … Butterworth, A. S. (2018). Genomic atlas of the human plasma proteome. Nature, 558(7708), 73–79. doi:10.1038/s41586-018-0175-2

t, K., Lahtela E, Havulinna AS, & Consortium., F. (2020). https://www.finngen.fi/en/researchers/clinical-endpoints.

Tran, T., Yang, J., Gardner, J., & Xiong, Y. (2018). GDF15 deficiency promotes high fat diet-induced obesity in mice. PLoS One, 13(8), e0201584. doi:10.1371/journal.pone.0201584

Tsai, V. W., Macia, L., Feinle-Bisset, C., Manandhar, R., Astrup, A., Raben, A., … Breit, S. N. (2015). Serum Levels of Human MIC-1/GDF15 Vary in a Diurnal Pattern, Do Not Display a Profile Suggestive of a Satiety Factor and Are Related to BMI. PLoS One, 10(7), e0133362. doi:10.1371/journal.pone.0133362

Tsai, V. W. W., Husaini, Y., Sainsbury, A., Brown, D. A., & Breit, S. N. (2018). The MIC-1/GDF15-GFRAL Pathway in Energy Homeostasis: Implications for Obesity, Cachexia, and Other Associated Diseases. Cell Metab, 28(3), 353–368. doi:10.1016/j.cmet.2018.07.018

Vartiainen, E., Jousilahti, P., Alfthan, G., Sundvall, J., Pietinen, P., & Puska, P. (2000). Cardiovascular risk factor changes in Finland, 1972-1997. Int J Epidemiol, 29(1), 49–56.

Verbanck, M., Chen, C. Y., Neale, B., & Do, R. (2018). Detection of widespread horizontal pleiotropy in causal relationships inferred from Mendelian randomization between complex traits and diseases. Nat Genet, 50(5), 693–698. doi:10.1038/s41588-018-0099-7

Vila, G., Riedl, M., Anderwald, C., Resl, M., Handisurya, A., Clodi, M., … Luger, A. (2011). The relationship between insulin resistance and the cardiovascular biomarker growth differentiation factor-15 in obese patients. Clin Chem, 57(2), 309–316. doi:10.1373/clinchem.2010.153726

Wang, D., Day, E. A., Townsend, L. K., Djordjevic, D., Jorgensen, S. B., & Steinberg, G. R. (2021). GDF15: emerging biology and therapeutic applications for obesity and cardiometabolic disease. Nat Rev Endocrinol, 17(10), 592–607. doi:10.1038/s41574-021-00529-7

Wiklund, F. E., Bennet, A. M., Magnusson, P. K., Eriksson, U. K., Lindmark, F., Wu, L., … Brown, D. A. (2010). Macrophage inhibitory cytokine-1 (MIC-1/GDF15): a new marker of all-cause mortality. Aging Cell, 9(6), 1057–1064. doi:10.1111/j.1474-9726.2010.00629.x

Wilding, J. P. H., Batterham, R. L., Calanna, S., Davies, M., Van Gaal, L. F., Lingvay, I., … Group, S. S. (2021). Once-Weekly Semaglutide in Adults with Overweight or Obesity. N Engl J Med, 384(11), 989. doi:10.1056/NEJMoa2032183

Willer, C. J., Li, Y., & Abecasis, G. R. (2010). METAL: fast and efficient meta-analysis of genomewide association scans. Bioinformatics, 26(17), 2190–2191. doi:10.1093/bioinformatics/btq340

Wollert, K. C., Kempf, T., Giannitsis, E., Bertsch, T., Braun, S. L., Maier, H., … Christenson, R. H. (2017). An Automated Assay for Growth Differentiation Factor 15. J Appl Lab Med, 1(5), 510–521. doi:10.1373/jalm.2016.022376

Wollert, K. C., Kempf, T., Lagerqvist, B., Lindahl, B., Olofsson, S., Allhoff, T., … Wallentin, L. (2007). Growth differentiation factor 15 for risk stratification and selection of an invasive treatment strategy in non ST-elevation acute coronary syndrome. Circulation, 116(14), 1540–1548. doi:10.1161/CIRCULATIONAHA.107.697714

Xiong, Y., Walker, K., Min, X., Hale, C., Tran, T., Komorowski, R., … Veniant, M. M. (2017). Long-acting MIC-1/GDF15 molecules to treat obesity: Evidence from mice to monkeys. Sci Transl Med, 9(412). doi:10.1126/scitranslmed.aan8732

Yang, L., Chang, C. C., Sun, Z., Madsen, D., Zhu, H., Padkjaer, S. B., … Jorgensen, S. B. (2017). GFRAL is the receptor for GDF15 and is required for the anti-obesity effects of the ligand. Nat Med, 23(10), 1158–1166. doi:10.1038/nm.4394

Yengo L, Sidorenko J, Kemper KE, Zheng Z, Wood AR, Weedon MN, … Consortium, G. (2018). Meta-analysis of genome-wide association studies for height and body mass index in ∼700,000 individuals of European ancestry. BioRxiv, doi: https://doi.org/10.1101/274654

Yumuk, V., Tsigos, C., Fried, M., Schindler, K., Busetto, L., Micic, D., … Obesity., O. M. T. F. o. t. E. A. f. t. S. o. (2016). European Guidelines for Obesity Management in Adults Obes Facts, 9(1), 64. doi:10.1159/000444869

Zimmers, T. A., Jin, X., Hsiao, E. C., McGrath, S. A., Esquela, A. F., & Koniaris, L. G. (2005). Growth differentiation factor-15/macrophage inhibitory cytokine-1 induction after kidney and lung injury. Shock, 23(6), 543–548.

