## Supplementary Materials for "Integrated Analyses of Growth Differentiation Factor-15 Concentration and Cardiometabolic Diseases in Humans"

**This document contains the supplementary materials for the manuscript by Lemmelä et al: "Integrated Analyses of Genetic Determinants of Growth Differentiation Factor-15 Concentration as a Cause for Metabolic Syndrome in Humans ".**

### Supplementary Methods

#### Study phenotypes

*FINRISK*

A wealth of quantitative biomarkers (e.g. blood lipids, blood sugar markers, inflammatory biomarkers, cytokines, blood count, fatty acids, metabolome) and physiological measures (e.g. anthropometrics, cardiovascular physiology, body composition) were collected from the study participants. In addition, participants who consented were matched to their electronic health records giving access to hospital discharge registry (years 1969-2015), hospital discharge registry of specialist health care operations (1996-2015), death registry (1992-2015), drug re-imbursement registry (1964-2015), drug purchase registry (1995-2015) and cancer registry (1953-2014). Gender was obtained from social security numbers.

*INTERVAL*

Participants were generally healthy because people with a history of major diseases (such as myocardial infarction, stroke, cancer, HIV, and hepatitis B or C) and those who have had recent illness or infection were ineligible to donate blood. Participants completed an online questionnaire which included questions about demographic characteristics (for example, age, sex and ethnicity), anthropometry measures (height, weight), and lifestyle information (alcohol intake, smoking, physical activity and diet)

#### Genotyping and imputation

*FINRISK*

26,404 FINRISK samples were genotyped using several arrays: the HumanCoreExome BeadChip, the Human610-Quad BeadChip, the Affymetrix6.0, and the Infinium

HumanOmniExpress (Illumina Inc., San Diego, CA, USA). The present study, using samples taken in FINRISK in 1997, consisted of 6,538 individuals which were genotyped using three genotyping arrays: the HumanCoreExome BeadChip, the Human610-Quad BeadChip, and the Infinium HumanOmniExpress (Illumina Inc., San Diego, CA, USA). Genotype calls were generated together with other available data sets using zCall at the Institute for Molecular Medicine Finland (FIMM). After sample-wise quality control (exclude samples with ambiguous gender, missingness (>5%), excess heterozygosity (±4sd), non-European ancestry) and variant-wise quality control (exclude SNPs with high missingness (>2%), low HWE P-value (<1e-6), minor allele count (MAC) <3 (in case Zcall'ed chip data) or MAC <10 (chip data called using Illumina GenCall) steps, the samples were pre-phased using Eagle2 (version 2.3). Genotype imputation was carried out by using a Finnish population-specific reference panel consisting of 2690 high-coverage WGS and 5092 WES samples with IMPUTE2 (version 2.3.2) (1) that allows the usage of two panels at the same time (the 'merge_ref_panels' option). Post-imputation quality control involved excluding variants imputed with imputation INFO < 0.7.

*INTERVAL*

The genotyping protocol and QC for the INTERVAL samples have previously been described in detail (2). DNA was extracted from buffy coat at LGC Genomics (UK) and was used to assay approximately 830,000 variants on the UK Biobank Affymetrix Axiom genotyping array at Affymetrix (Santa Clara, California, US). Genotyping was performed in batches of approximately 4,800 samples. Variants were excluded from a batch if they strongly deviated from HWE (p value <5e-6) or had a within-batch call rate <0.97. Sample QC included removing duplicate samples and samples with non-European ancestry, missing phenotypic sex and sex mismatches and extreme heterozygosity (±3SD). Relatedness was removed by excluding one participant from each pair of close (first- or second-degree) relatives, defined as $\hat{\pi} > 0.187$.

Additional variant QC steps were performed prior to imputation to establish a high quality imputation scaffold. This included imposing a global HWE filter of p value <5e-6, a call rate filter of 99% over the INTERVAL genotyping batches that a variant was not failed in, and a global call rate filter of 75% across all INTERVAL genotyping batches. All monomorphic variants, non-autosomal and multi-allelic variants were removed and 654,966 high-quality variants remained to be used for imputation. Phasing was conducted using SHAPEIT3 and variants were imputed using a combined 1000 Genomes Phase 3-UK10K imputation panel. Imputation was performed on the Sanger Imputation Server (https://imputation.sanger.ac.uk), resulting in 87,696,888 imputed variants.

#### UK Biobank exome sequencing

UK Biobank was whole exome sequenced (paired-end 75bp) at Regeneron Pharmaceuticals using the IDT xGen v1 capture kit and NovaSeq6000 (for more information see previous publication (3)). Data was available on 302,362 individuals of which >95% of CCDS had at least 10x coverage and average coverage of CCDS was 59X. Alignment to GRCh38 and SNV and indel calling were completed utilising Illumina DRAGEN Bio-IT Platform Germline Pipeline v3.07 on a custom-built Amazon Web Services (AWS) cloud platform. Annotation of SNVs and indels was performed using SnpEFF v4.3 against Ensembl Build 38.92 and MAPQ < 30 were excluded. Related individuals were identified using KING and first and second degree relatives were excluded from downstream analysis. Sex and ancestry checks were completed using PEDDY (4) and individuals with gender mismatch were excluded.

#### Variant annotation

The functional significance of the fine mapped variants were explored using public databases and browsers, including *GTEx, Ensmbl, SNiPA, RegulomeDB* (5).

#### Mendelian Randomisation

In brief, Mendelian randomisation is a method that explores whether an intermediate trait has a causal relationship with an endpoint by using genetic instruments. The use of genetic data means there is less environmental bias. Variants identified as significantly associated with the intermediate trait are applied and summary results from intermediate trait – variants and endpoint – variants are applied in the model. MR-Egger regression does not constrain the intercept, therefore it is not biased by invalid IV but has reduced power.

### Supplementary Results

#### Genome-wide association of GDF15 levels

*FINRISK*

In FINRISK GWAS was conducted on 5,817 individuals with available GDF15 plasma levels, measured using an immunoluminometric assay (6) (Supplementary Table S8). The most significant associations were found within or in proximity to the *GDF15* gene on chromosome 19, comprising 159 significant variants ($p$-value<$5x10^{-8}$). Statistical fine mapping analysis (using FINEMAP (7)) within the *GDF15* gene region revealed four independent putative causal variants with a probability of 0.85. This configuration included rs16982345, rs1054221, rs1059369 and rs189593084. The regional heritability of these four fine mapped variants explained 10% (95% CI: 8-12%) of the GDF15 variation whereas the signal driven by the variant rs16982345 explained 6% of the variation alone. In addition to this locus, two genome-wide significant rare variants were identified on chromosome 7 in the *CAPZA2* gene; rs200430819 (MAF=0.001) and rs200765554 (MAF=0.001).

*INTERVAL*

In INTERVAL we assayed plasma GDF15 in 3,301 participants using SOMAScan (an aptamer-based multiplex protein assays) and in 4,998 participants using a proximity extension-based antibody assay (Olink).

The most significant associations were found around the *GDF15* gene on chromosome 19, comprising 134 significant variants for INTERVAL-SOMAscan and 72 significant variants for INTERVAL-Olink ($p$-value<$5x10^{-8}$) (Supplementary Tables S9-10).

**Fine mapped variants in independent studies from unconditioned GWAS**

*FINRISK*

Statistical fine mapping analysis (using FINEMAP (7)) of this locus revealed four putative causal variants with a probability of 0.85. This configuration included rs16982345, rs1054221, rs1059369 and rs189593084. The regional heritability of these four fine mapped variants explained 10% (95% CI: 8-12%) of the GDF15 variation whereas the signal driven by the variant rs16982345 explained 6% of the variation alone. In addition to this locus, two genome-wide significant rare variants were identified on chromosome 7 in the *CAPZA2* gene; rs200430819 (MAF=0.001) and rs200765554 (MAF=0.001).

*INTERVAL*

Statistical fine mapping analysis in INTERVAL-SomaScan of this locus using FINEMAP revealed three putative causal variants with a probability of 0.885 for INTERVAL-SOMAscan and the top configuration included rs1227734, rs1058587 and rs3787023. The regional heritability of these three fine mapped variants explained 20.2% (95% CI: 17.8-23.0%) of the GDF15 variation.

Statistical fine mapping analysis in INTEVAL-Olink of this locus revealed three putative causal variants with a probability of 0.632. However, the top configuration identified by FINEMAP only included two genetic variants, rs1054221 and rs1055150 and had a regional heritability of 7.6% (95% CI: 6.2-9.0%).

### Supplementary Tables and Figures

**Supplementary Table S1. FINRISK cohort characteristics.**

| | **Overall (N= 6610)** | **Female (N=3257)** | **Male (N= 3353)** |
|---|---|---|---|
| *GDF15 (ng/L, mean, ± SD, range)* | 1053 (2046) | 1166 (2858) | 943 (542) |
| *Baseline age (years, mean, ± SD, range)* | 47.7 (13.1) | 46.2 (12.6) | 49.2 (13.4) |
| **Cardiometabolic risk factors** | | | |
| *Smoking (%, n)* | 26.1 (6555) | 21.0 (3237) | 31.1 (3318) |
| *Body mass Index (kg/m$^2$; mean, ± SD, range, n)* | 26.6 (4.5, 6602) | 25.3 (5.0, 3254) | 27.0 (4.0, 3348) |
| *Systolic Blood Pressure (mmHg; mean, ± SD, range, n)* | 136 (19, 6604) | 131 (19, 3254) | 140 (19, 3350) |
| *Total Cholesterol (mmol/L; mean, ± SD, range)* | 5.5 (1.1) | 5.5 (1.1) | 5.6 (1.0) |
| *Triglycerides (mmol/L; mean, ± SD, range)* | 1.5 (1.1) | 1.3 (0.9) | 1.7 (1.1) |
| | **Overall (N=6600)** | **Female (N=3248)** | **Male (N= 3352)** |
| **Outcome** | | | |
| *All-cause mortality (%)* | 16.4 | 10.3 | 22.3 |
| *Type 2 Diabetes (%)* | 8.9 | 6.7 | 10.5 |
| *Myocardial infarction (%)* | 7.2 | 3.7 | 10.5 |
| *Heart Failure (%)* | 6.2 | 4.4 | 8.0 |
| *Cardiovascular disease (%)* | 51.7 | 50.1 | 52.1 |

**Supplementary Table S2. Baseline characteristics in FINRISK associated with GDF15 plasma levels.**

| | Beta (SE) | *p*-value | Variance of GDF15 explained (%) |
|---|---|---|---|
| **Age** | 0.041 (0.00080) | $< 2.2 \times 10^{-308}$ | 28.0 |
| **Gender** | -0.033 (se=0.021) | 0.12 | - |
| **BMI** | 0.039 (se=0.011) | 0.00051 | 0.1 |
| **Smoking** | 0.34 (0.024) | $2.2 \times 10^{-44}$ | 1.9 |

**Supplementary Table S3. GDF15 level disease associations corrected for age and sex only.** Abbreviations: SAH, aneurysmal subarachnoid haemorrhage; ANGIO, coronary angioplasty; CABG, coronary artery bypass grafting; DVT, deep vein thrombosis – in separate excel file

**Supplementary Table S4. GDF15 level quantitative biomarker associations corrected for age and sex only.** All quantitave biomarkers were rank-based inverse transformed – in separate excel file

**Supplementary Table S5. GDF15 level disease associations corrected for age, sex, BMI and smoking**– in separate excel file

**Supplementary Table S6. Independent predictors of all-cause mortality, Type 2 diabetes and Cardiovascular disease (CHD or STR) event.**

| Predictors for all-cause mortality | | | |
|---|---|---|---|
| | **HR** | **95%CI** | **P-value** |
| **GDF15** | **1.66** | **1.51 – 1.80** | **$9.19x10^{-12}$** |
| Mean systolic blood pressure (mmHg) | 1.01 | 0.90 -1.12 | 0.81 |
| Blood pressure lowering medication | 1.16 | 0.92 – 1.40 | 0.23 |
| Total cholesterol (mmol/l) | 0.99 | 0.88 – 1.10 | 0.86 |
| Cancers, any | 2.35 | 2.14 – 2.56 | $5.94x10^{-16}$ |
| HDL cholesterol (mmol/l) | 0.92 | 0.81 – 1.04 | 0.17 |
| Smoking (yes/no) | 1.93 | 1.71 – 2.15 | $6.48x10^{-9}$ |
| Prevalent Cardiovascular diseases (CHD or STR) | 1.91 | 1.59 – 2.23 | $8.39x10^{-5}$ |
| Prevalent Diabetes (type II or I) | 1.25 | 0.95 – 1.54 | 0.14 |
| Obesity (BMI>30) | 1.12 | 0.88 – 1.36 | 0.34 |

| Predictors for Cardiovascular disease (CHD or STR) | | | |
|---|---|---|---|
| **GDF15** | **1.39** | **1.26 – 1.52** | **$1.50x10^{-6}$** |
| Mean systolic blood pressure (mmHg) | 1.11 | 1.00 – 1.22 | 0.07 |
| Blood pressure lowering medication | 1.07 | 0.84 – 1.30 | 0.57 |
| Total cholesterol (mmol/l) | 1.15 | 1.05 – 1.26 | 0.008 |
| HDL cholesterol (mmol/l) | 0.70 | 0.59 – 0.81 | $1.07x10^{-10}$ |
| Smoking (yes/no) | 1.39 | 1.18 – 1.61 | 0.003 |
| Prevalent Diabetes (type II or I) | 1.62 | 1.35 – 1.89 | 0.0005 |
| Lipid medication | 1.45 | 1.08 – 1.82 | 0.05 |
| Obesity (BMI>30) | 1.13 | 0.91 – 1.35 | 0.26 |

| Predictors for type II diabetes | | | |
|---|---|---|---|
| **GDF15** | **1.40** | 1.12 – 1.68 | **0.02** |
| Mean systolic blood pressure (mmHg) | 1.03 | 0.80 – 1.26 | 0.78 |
| Blood pressure lowering medication | 1.36 | 0.90 – 1.82 | 0.19 |
| Total cholesterol (mmol/l) | 1.21 | 1.00 – 1.43 | 0.07 |
| HDL cholesterol (mmol/l) | 0.52 | 0.28 – 0.76 | $1.30x10^{-7}$ |
| Smoking (yes/no) | 1.56 | 1.09 – 2.03 | 0.06 |
| Prevalent Cardiovascular diseases (CHD or STR) | 1.03 | 0.21 – 1.84 | 0.95 |
| Obesity (BMI>30) | 3.78 | 3.35 – 4.22 | $2.75x10^{-9}$ |

Cox proportional hazard model was used to estimate the associations between risk factors and outcomes.

**Supplementary Table S7. GDF15 level biomarker associations corrected for age, sex, smoking and BMI** All quantitave biomarkers were rank-based inverse transformed – in separate excel file

**Supplementary Table S8. Significant (*p*-value < $5x10^{-8}$) genome-wide association study for FINRISK** – in separate excel file

**Supplementary Table S9. Significant (*p*-value < $5x10^{-8}$) genome-wide association study for Somalogic** – in separate excel file

**Supplementary Table S10. Significant (*p*-value < $5x10^{-8}$) genome-wide association study for Olink** – in separate excel file

**Supplementary Table S11. Meta-analysis of FINRISK and INTERVAL GDF15 GWAS results.**

| | | | **FINRISK** | | **INTERVAL-SomaScan** | | **INTERVAL-Olink** | | **Meta-analysis** | | | |
|---|---|---|---|---|---|---|---|---|---|---|---|---|
| **SNPs** | **LD block** | **EA/OA** | **beta** | ***p*-value** | **beta** | ***p*-value** | **beta** | ***p*-value** | **beta** | ***p*-value** | **Heterogeneity $I^2$** | **Heterogeneity *p*-value** |
| rs16982345 | 1 | A/G | -0.34 | **$4.6x10^{-83}$** | 0.57 | **$1.1x10^{-92}$** | 0.01 | 0.72 | -0.05 | $3.7x10^{-5}$ | 99.8% | **$3.2x10^{-177}$** |
| rs1058587 | 1 | G/C | -0.34 | **$5.5x10^{-83}$** | 0.57 | **$1.3x10^{-92}$** | 0.01 | 0.77 | -0.05 | $3.4x10^{-5}$ | 99.8% | **$5.4x10^{-177}$** |
| rs3787023 | 1 | A/G | -0.11 | **$8.9x10^{-12}$** | 0.29 | **$7.0x10^{-33}$** | 0.04 | 0.072 | 0.02 | 0.066 | 99.0% | **$1.6x10^{-42}$** |
| rs1055150 | 1 | G/C | -0.11 | **$8.3x10^{-12}$** | 0.29 | **$3.3x10^{-32}$** | 0.04 | 0.080 | 0.02 | 0.086 | 98.9% | **$6.9x10^{-42}$** |
| rs1059369 | 1 | A/T | 0.18 | **$8.2x10^{-28}$** | -0.19 | **$1.0x10^{-10}$** | 0.04 | 0.078 | 0.09 | **$2.0x10^{-10}$** | 98.4% | **$5.9x10^{-28}$** |
| rs1054221 | 2 | C/T | 0.48 | **$1.1x10^{-54}$** | 0.41 | **$1.7x10^{-32}$** | 0.52 | **$9.4x10^{-74}$** | 0.48 | **$3.5x10^{-160}$** | 64.3% | 0.061 |
| rs1227734 | 2 | T/C | 0.48 | **$1.3x10^{-54}$** | 0.42 | **$7.1x10^{-34}$** | 0.51 | **$7.1x10^{-74}$** | 0.47 | **$7.6x10^{-162}$** | 58.7% | 0.089 |
| rs189593084 | 3 | A/C | -0.25 | $6.4x10^{-8}$ | - | - | -0.47 | 0.0053 | -0.26 | **$2.5x10^{-9}$** | 38.0% | 0.20 |

Variants listed in table were identified by fine mapping GWAS results from FINRISK and INTERVAL. LD blocks were defined as SNPs that had LD > 0.1 with the lead variant (most significantly associated variant).

**Supplementary Table S12. Linkage disequilibrium ($R^2$) between fine mapped variants in FINRISK, Somalogic and Olink.**

| | rs16982345 | rs1054221 | rs1059369 | rs189593084 | rs3787023 | rs1058587 | rs1227734 | rs1055150 |
|---|---|---|---|---|---|---|---|---|
| **rs16982345** | | - | 0.11 | - | 0.34 | 1.0 | - | 0.33 |
| **rs1054221** | - | | - | - | 0.16 | - | 1.0 | 0.16 |
| **rs1059369** | 0.11 | - | | - | 0.33 | 0.11 | - | 0.33 |
| **rs189593084** | - | - | - | | - | - | - | - |
| **rs3787023** | 0.34 | 0.16 | 0.33 | - | | 0.34 | 0.16 | 0.99 |
| **rs1058587** | 1.0 | - | 0.11 | - | 0.34 | | - | 0.33 |
| **rs1227734** | - | 1.0 | - | - | 0.16 | - | | 0.16 |
| **rs1055150** | 0.33 | 0.16 | 0.33 | - | 0.99 | 0.33 | 0.16 | |

**Supplementary Table S13. GDF15 level disease associations conditioned on rs1058587 as well as corrected for age, sex, smoking and BMI** – in separate excel file
**Supplementary Table S14. GDF15 level biomarker associations conditioned on rs1058587 as well as corrected for age, sex, smoking and BMI** – in separate excel file
**Supplementary Table S15. Significant (*p*-value < $5x10^{-8}$) genome-wide association study conditioned for rs1058587 in FINRISK** – in separate excel file
**Supplementary Table S16. Significant (*p*-value < $5x10^{-8}$) genome-wide association study meta-analysis conditioned for rs1058587 in INTERVAL-SomaScan** – in separate excel file
**Supplementary Table S17. Significant (*p*-value < $5x10^{-8}$) genome-wide association study meta-analysis conditioned for rs1058587 in FINRISK, INTERVAL-SomaScan and INTERVAL-Olink** – in separate excel file

**Supplementary Table S18. Functional annotation of GDF15 meta-analysis fine mapped variants.**

| Variant | Freq | Allele | Gene | Consequence | Biotype |
|---|---|---|---|---|---|
| rs1054221 | 0.10 | C | GDF15 | 3 prime UTR variant | protein_coding |
| | | C | LRRC25 | Downstream gene variant | protein_coding |
| | | C | MIR3189 | Downstream gene variant | miRNA |
| rs3787023 | 0.43 | A or C | GDF15 | Intron | protein_coding |
| | | A or C | LRRC25 | Downstream gene variant | protein_coding |
| | | A or C | MIR3189 | Downstream gene variant | miRNA |
| rs138515339 | <0.01 | A or T | CRTC1 | Intron | protein_coding |
| rs141542836 | 0.02 | T | GDF15 | 5 prime UTR variant | protein_coding |
| | | T | LRRC25 | Downstream gene variant | protein_coding |
| | | T | MIR3189 | Downtream gene variant | miRNA |

Canonical transcripts only are shown. Results were obtained from Ensembl variant effect predictor. Frequencies were obtained from 1000 genomes.

**Supplementary Table S19. GDF15 PTV carrier frequency in UKB.**

| Mutation | Variant | Total Freq | EUR Freq | AFR Freq | SAS Freq |
|---|---|---|---|---|---|
| A189-R190 frameshift | 19-18388575-CA-C | 0.50 | 0.55 | 0 | 0.13 |
| Q187 stop gained | 19-18388567-C-T | 0.06 | 0.02 | 0 | 0.75 |
| L30-S31 frameshift | 19-18386279-GTC-G | 0.05 | 0.05 | 0 | 0 |
| S36 frameshift | 19-18386297-C-CCCGGGACCCT | 0.05 | 0 | 1.0 | 0 |
| V93 splice donor | 19-18386468-T-C | 0.04 | 0.04 | 0 | 0 |
| H26 frameshift | 19-18386267-T-TG | 0.04 | 0.04 | 0 | 0 |
| M1 start lost | 19-18386192-G-T | 0.01 | 0.01 | 0 | 0 |
| S44 stop gained | 19-18386320-C-A | 0.01 | 0.01 | 0 | 0 |
| R54 stop gained | 19-18386349-C-T | 0.01 | 0 | 0 | 0.13 |
| K58 stop gained | 19-18386361-A-T | 0.03 | 0.03 | 0 | 0 |
| Y60 stop gained | 19-18386369-C-G | 0.01 | 0.01 | 0 | 0 |
| E74 stop gained | 19-18386409-G-T | 0.01 | 0.01 | 0 | 0 |
| V93 splice acceptor | 19-18388285-G-A | 0.01 | 0.01 | 0 | 0 |
| W135 stop gained | 19-18388412-G-A | 0.01 | 0 | 0 | 0.13 |
| W225 stop gained | 19-18388683-G-A | 0.01 | 0.01 | 0 | 0 |
| W228 stop gained | 19-18388691-G-A | 0.01 | 0.01 | 0 | 0 |
| S231 stop gained | 19-18388700-C-A | 0.01 | 0.01 | 0 | 0 |
| Q266 stop gained | 19-18388864-C-T | 0.01 | 0.01 | 0 | 0 |
| A33 frameshift | 19-18386287-C-CTCGCATG | 0.01 | 0.01 | 0 | 0 |
| S44 frameshift | 19-18386320-C-CAGAA | 0.01 | 0.01 | 0 | 0 |
| S76-T78 frameshift | 19-18386415-TCGAACAC-T | 0.01 | 0.01 | 0 | 0 |
| I88-L89 frameshift | 19-18386452-TACTC-T | 0.03 | 0.03 | 0 | 0 |
| R149-F150 frameshift | 19-18388455-AC-A | 0.03 | 0.03 | 0 | 0 |
| G191-A195 frameshift | 19-18388580-GGCGCCGCAGA-G | 0.02 | 0.02 | 0 | 0 |
| D201-P204 frameshift | 19-18388610-ACCACTGT-A | 0.01 | 0.01 | 0 | 0 |
| H202-C203 frameshift | 19-18388614-CT-C | 0.01 | 0.01 | 0 | 0 |
| C210-C211 frameshift | 19-18388638-CT-C | 0.02 | 0.02 | 0 | 0 |
| T290-T290 frameshift | 19-18388876-ACC-A | 0.01 | 0.01 | 0 | 0 |

### Supplementary Figures

### Supplementary Figure S1. GDF15 plasma levels in FINRISK.

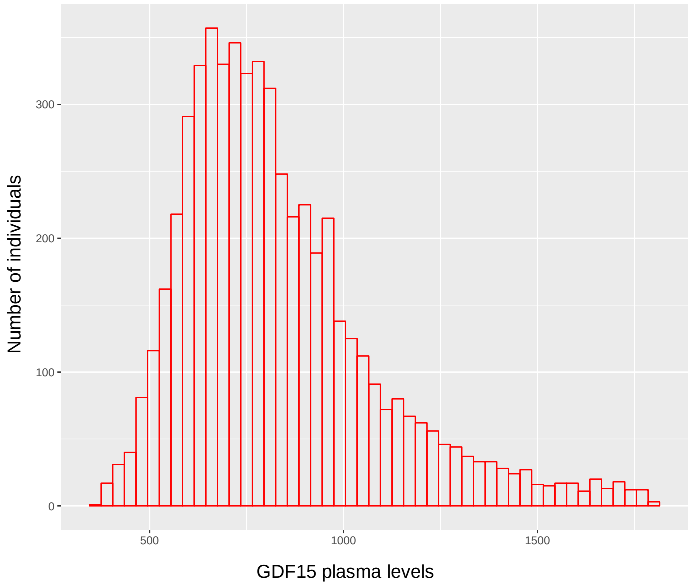


Histogram showing individuals with levels ≥ 1800ng/l removed

**Supplementary Figure S2. GDF15 plasma concentration in prevalent and incident type 2 diabetes cases and healthy controls.**

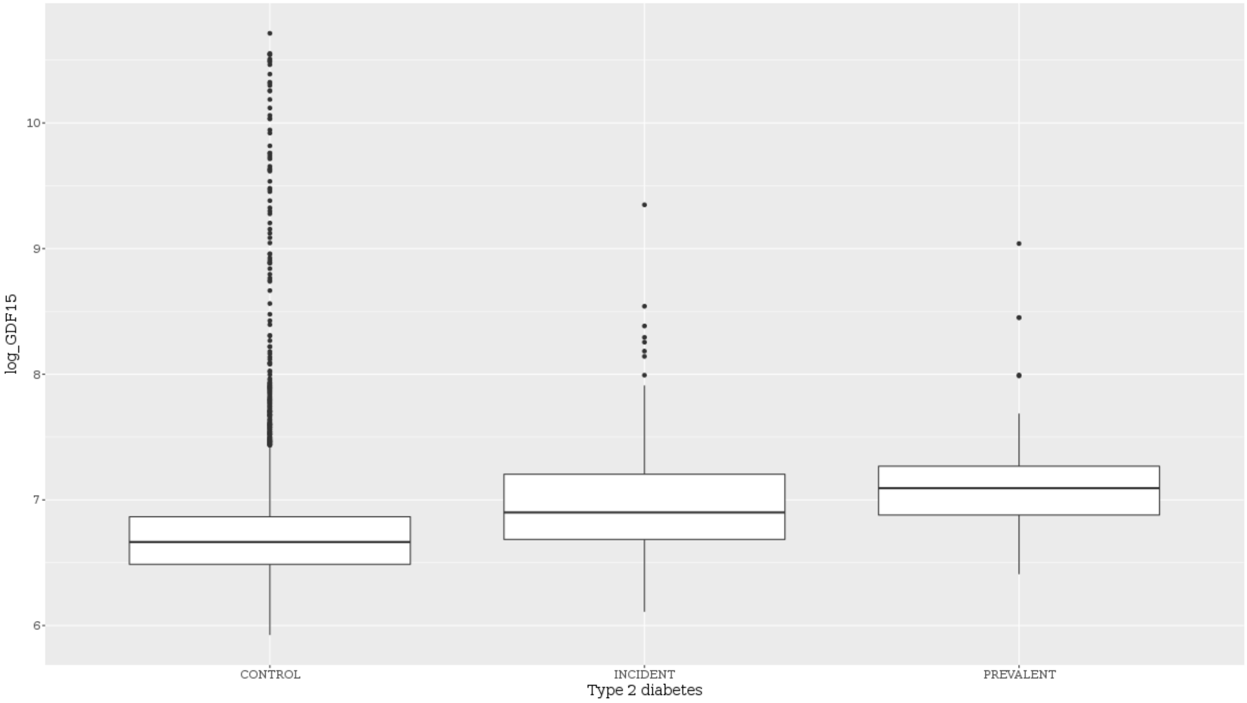


Prevalent cases were type II diabetic patients diagnosed before levels were taken, incident cases were were diagnosed after GDF15 levels were taken, and controls did not have a dignosis of diabetes.

**Supplementary Table S3. Survival curves for cox proportional hazard models on ten years follow-up according to GDF15 quartiles.**

a)

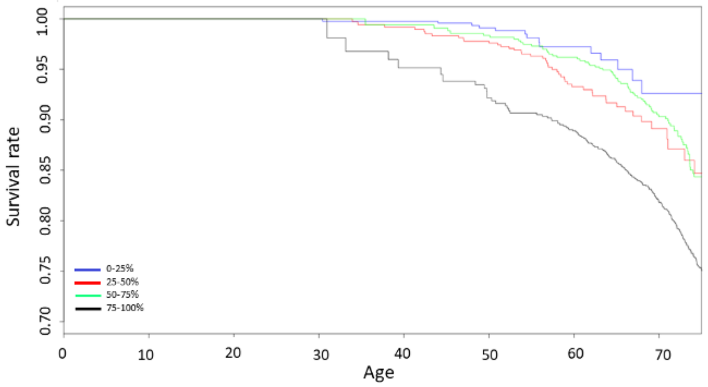


b)

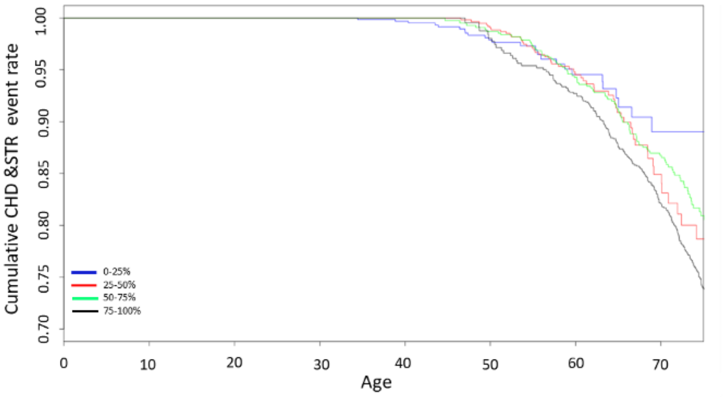


c)

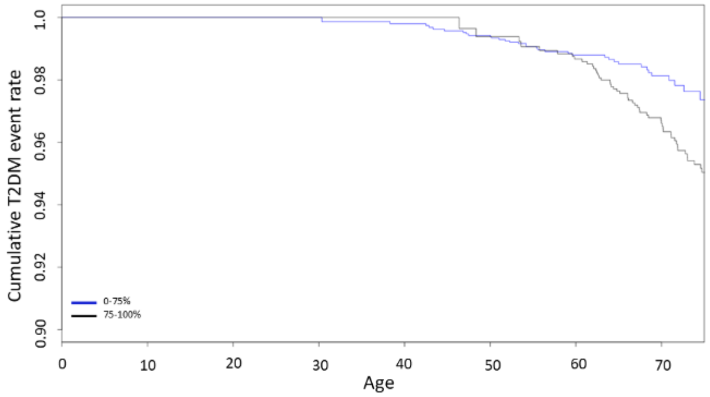


Figures represent survival curves in (a) death, (b) cardiovascular disease and (c) diabetes and are divided into quantiles. Type 2 diabetes shows only a comparison of the last quartile (75-100%) due to insufficient power when treating the other quartiles separately. Abbreviations: T2DM, type 2 diabetes mellitus.

**Supplementary Figure S4. Manhattan and Q-Q plot for the GWAS meta-analysis of conditioned GDF15 plasma levels in 14,099 individuals.**

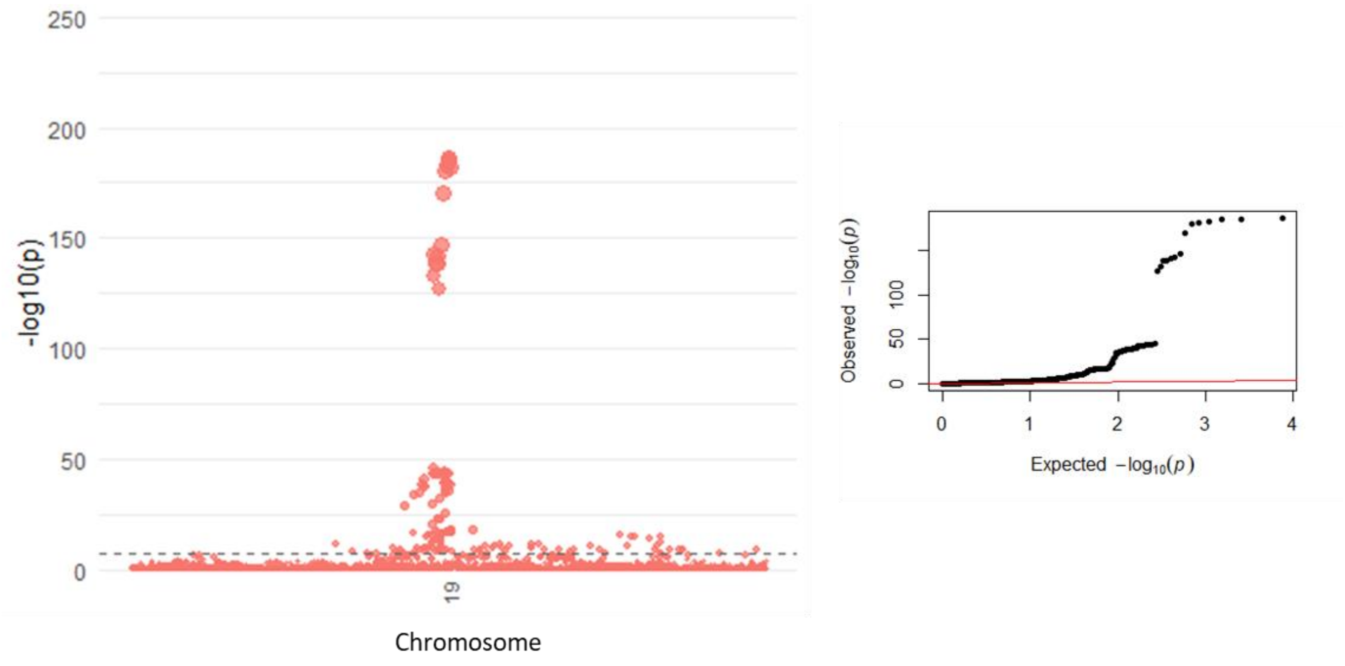


The black line represents genome-wide significance ($p$-value<$5x10^{-8}$).

**Supplementary Figure S5. Mendelian randomisation graphs for GDF15 as exposure and outcomes as (a) BMI, (b) HDL and (c) BMD**

**a)**

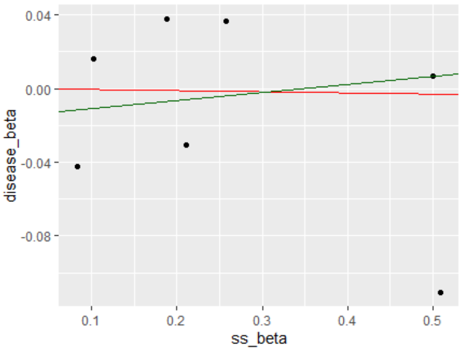


**b)**

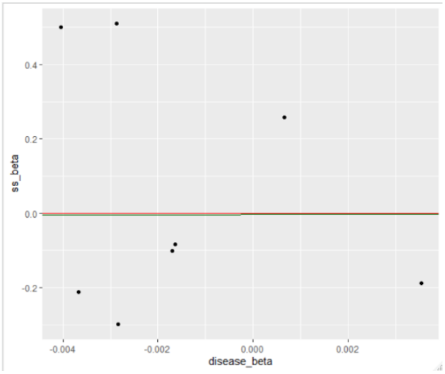


**c)**

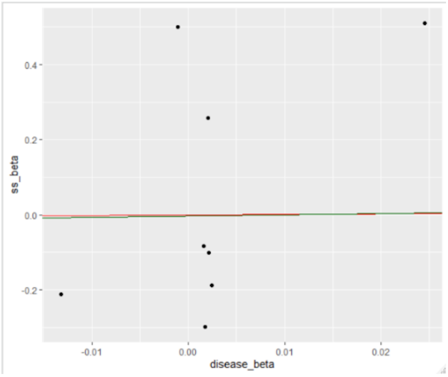


No significant causal association was found with any outcome. Axis represent the estimated effect from the GWAS summary statistics, the red line represents the MR IVW line of fit and the green line represents the MR-Egger line of fit.

**Supplementary Figure S6. Forest plot demonstrating heterogeneity between rs1058587 in FINRISK and INTERVAL.**

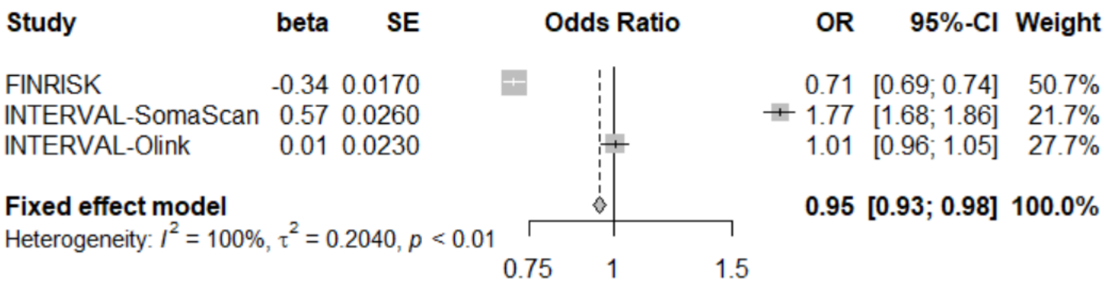

### References


1. Howie BN, Donnelly P, Marchini J. A flexible and accurate genotype imputation method for the next generation of genome-wide association studies. PLoS Genet. 2009;5(6):e1000529.
2. Astle WJ, Elding H, Jiang T, Allen D, Ruklisa D, Mann AL, et al. The Allelic Landscape of Human Blood Cell Trait Variation and Links to Common Complex Disease. Cell. 2016;167(5):1415-29 e19.
3. Wang Q, Dhindsa RS, Carss K, Harper A, Nag A, Tachmazidou I, et al. Surveying the contribution of rare variants to the genetic architecture of human disease through exome sequencing of 177,882 UK Biobank participants. bioRxiv 2020:doi: https://doi.org/10.1101/2020.12.13.422582
4. Pedersen BS, Quinlan AR. Who's Who? Detecting and Resolving Sample Anomalies in Human DNA Sequencing Studies with Peddy. Am J Hum Genet. 2017;100(3):406-13.
5. Urmo Võsa AC, Harm-Jan Westra, Marc Jan Bonder, Patrick Deelen, Biao Zeng, Holger Kirsten, Ashis Saha, Roman Kreuzhuber, Silva Kasela, Natalia Pervjakova, Isabel Alvaes, Marie-Julie Fave, Mawusse Agbessi, Mark Christiansen, Rick Jansen, Ilkka Seppälä, Lin Tong, Alexander Teumer, Katharina Schramm, Gibran Hemani, Joost Verlouw, Hanieh Yaghootkar, Reyhan Sönmez, Andrew Brown, Viktorija Kukushkina, Anette Kalnapenkis, Sina Rüeger, Eleonora Porcu, Jaanika Kronberg-Guzman, Johannes Kettunen, Joseph Powell, Bernett Lee, Futao Zhang, Wibowo Arindrarto, Frank Beutner, BIOS Consortium, Harm Brugge, i2QTL Consortium, Julia Dmitreva, Mahmoud Elansary, Benjamin P. Fairfax, Michel Georges, Bastiaan T. Heijmans, Mika Kähönen, Yungil Kim, Julian C. Knight, Peter Kovacs, Knut Krohn, Shuang Li, Markus Loeffler, Urko M. Marigorta, Hailang Mei, Yukihide Momozawa, Martina Müller-Nurasyid, Matthias Nauck, Michel Nivard, Brenda Penninx, Jonathan Pritchard, Olli Raitakari, Olaf Rotzchke, Eline P. Slagboom, Coen D.A. Stehouwer, Michael Stumvoll, Patrick Sullivan, Peter A.C. ‘t Hoen, Joachim Thiery, Anke Tönjes, Jenny van Dongen, Maarten van Iterson, Jan Veldink, Uwe Völker, Cisca Wijmenga, Morris Swertz, Anand Andiappan, Grant W. Montgomery, Samuli Ripatti, Markus Perola, Zoltan Kutalik, Emmanouil Dermitzakis, Sven Bergmann, Timothy Frayling, Joyce van Meurs, Holger Prokisch, Habibul Ahsan, Brandon Pierce, Terho Lehtimäki, Dorret Boomsma, Bruce M. Psaty, Sina A. Gharib, Philip Awadalla, Lili Milani, Willem Ouwehand, Kate Downes, Oliver Stegle, Alexis Battle, Jian Yang, Peter M. Visscher, Markus Scholz, Gregory Gibson, Tõnu Esko, Lude Franke. Unraveling the polygenic architecture of complex traits using blood eQTL metaanalysis. BioRxiv. 2018:doi: https://doi.org/10.1101/447367
6. Sinning C, Ojeda F, Wild PS, Schnabel RB, Schwarzl M, Ohdah S, et al. Midregional proadrenomedullin and growth differentiation factor-15 are not influenced by obesity in heart failure patients. Clin Res Cardiol. 2017;106(6):401-10.
7. Benner C, Spencer CC, Havulinna AS, Salomaa V, Ripatti S, Pirinen M. FINEMAP: efficient variable selection using summary data from genome-wide association studies. Bioinformatics. 2016;32(10):1493-501.
